# Comparative gene regulatory networks modulating *APOE* expression in microglia and astrocytes

**DOI:** 10.1101/2024.04.19.24306098

**Authors:** Logan Brase, Yanbo Yu, Eric McDade, Dominantly Inherited Alzheimer Network (DIAN), Oscar Harari, Bruno A. Benitez

## Abstract

**Background:** Single-cell technologies have unveiled various transcriptional states in different brain cell types. Transcription factors (TFs) regulate the expression of related gene sets, thereby controlling these diverse expression states. Apolipoprotein E (*APOE*), a pivotal risk-modifying gene in Alzheimer’s disease (AD), is expressed in specific glial transcriptional states associated with AD. However, it is still unknown whether the upstream regulatory programs that modulate its expression are shared across brain cell types or specific to microglia and astrocytes.

**Methods:** We used pySCENIC to construct state-specific gene regulatory networks (GRNs) for resting and activated cell states within microglia and astrocytes based on single-nucleus RNA sequencing data from AD patients’ cortices from the Knight ADRC-DIAN cohort. We then identified replicating TF using data from the ROSMAP cohort. We identified sets of genes co-regulated with *APOE* by clustering the GRN target genes and identifying genes differentially expressed after the virtual knockout of TFs regulating *APOE*. We performed enrichment analyses on these gene sets and evaluated their overlap with genes found in AD GWAS loci.

**Results:** We identified an average of 96 replicating regulators for each microglial and astrocyte cell state. Our analysis identified the CEBP, JUN, FOS, and FOXO TF families as key regulators of microglial APOE expression. The steroid/thyroid hormone receptor families, including the THR TF family, consistently regulated APOE across astrocyte states, while CEBP and JUN TF families were also involved in resting astrocytes. AD GWAS-associated genes (*PGRN*, *FCGR3A*, *CTSH*, *ABCA1*, *MARCKS*, *CTSB*, *SQSTM1*, *TSC22D4*, *FCER1G*, and HLA genes) are co-regulated with APOE. We also uncovered that APOE-regulating TFs were linked to circadian rhythm (*BHLHE40*, *DBP*, *XBP1*, *CREM*, *SREBF1*, *FOXO3*, and *NR2F1*).

**Conclusions:** Our findings reveal a novel perspective on the transcriptional regulation of *APOE* in the human brain. We found a comprehensive and cell-type-specific regulatory landscape for *APOE*, revealing distinct and shared regulatory mechanisms across microglia and astrocytes, underscoring the complexity of *APOE* regulation. *APOE*-co-regulated genes might also affect AD risk. Furthermore, our study uncovers a potential link between circadian rhythm disruption and *APOE* regulation, shedding new light on the pathogenesis of AD.

## Introduction

Alzheimer disease (AD) is a progressive neurodegenerative disorder that affects millions of people worldwide. It is characterized by cognitive impairment, memory loss, and behavioral changes. The strongest genetic risk modifiers are variants in the gene *APOE*. It is involved in lipid transport and metabolism in the brain and has two primary variants that influence disease risk, e2 (protective) and e4 (risk). *APOE* is constitutively expressed in astrocytes and activated microglia^1^, cell types that are key players in neuroinflammation and neurodegenerative processes^2^.

When activated, microglial cells can significantly upregulate *APOE* expression, a process that is crucial for the phagocytic clearance of amyloid-beta and other debris^3^. Astrocytes, utilize *APOE* to support the lipid and energy demands of the surrounding neurons^4^. The ε4 allele, in particular, has been associated with a detrimental effect on microglial activation and astrocytic function, exacerbating neuroinflammatory and neurodegenerative processes^2–4^. These expression states are not static; they evolve in response to the brain’s pathological environment, influencing disease progression and severity. Understanding the differences in *APOE* expression regulation between microglial and astrocytic cell states can provide valuable insights into the molecular mechanisms underlying *APOE*‘s involvement in AD and potentially other neurodegenerative disorders.

In this study, we aimed to further investigate the differences in the regulation of *APOE* expression between various microglial and astrocytic cell states using single-nucleus RNA-seq (snRNA-seq). To this end, we performed state-specific gene regulatory network (GRN) analysis using pySCENIC on specific cell states (Figure 1). We then used two approaches to identify gene sets co-regulated with *APOE* in these networks: co-regulatory network clusters and transcription factor knockout (TF-KO) differentially expressed genes (DEGs). We assessed the relevance of these gene sets to AD by testing their overlap with AD genome-wide association study (GWAS) genes and their enrichment in AD-related pathways. By delineating the GRNs in these critical cell states, we seek to elucidate the distinct transcriptional regulatory mechanisms that govern *APOE* expression, which is crucial for unraveling the complex interplay between *APOE* regulation and AD pathogenesis. This knowledge may lead to the development of targeted therapeutic interventions to modulate *APOE*-related biology in specific cell types to mitigate disease progression.

**Figure 1.**
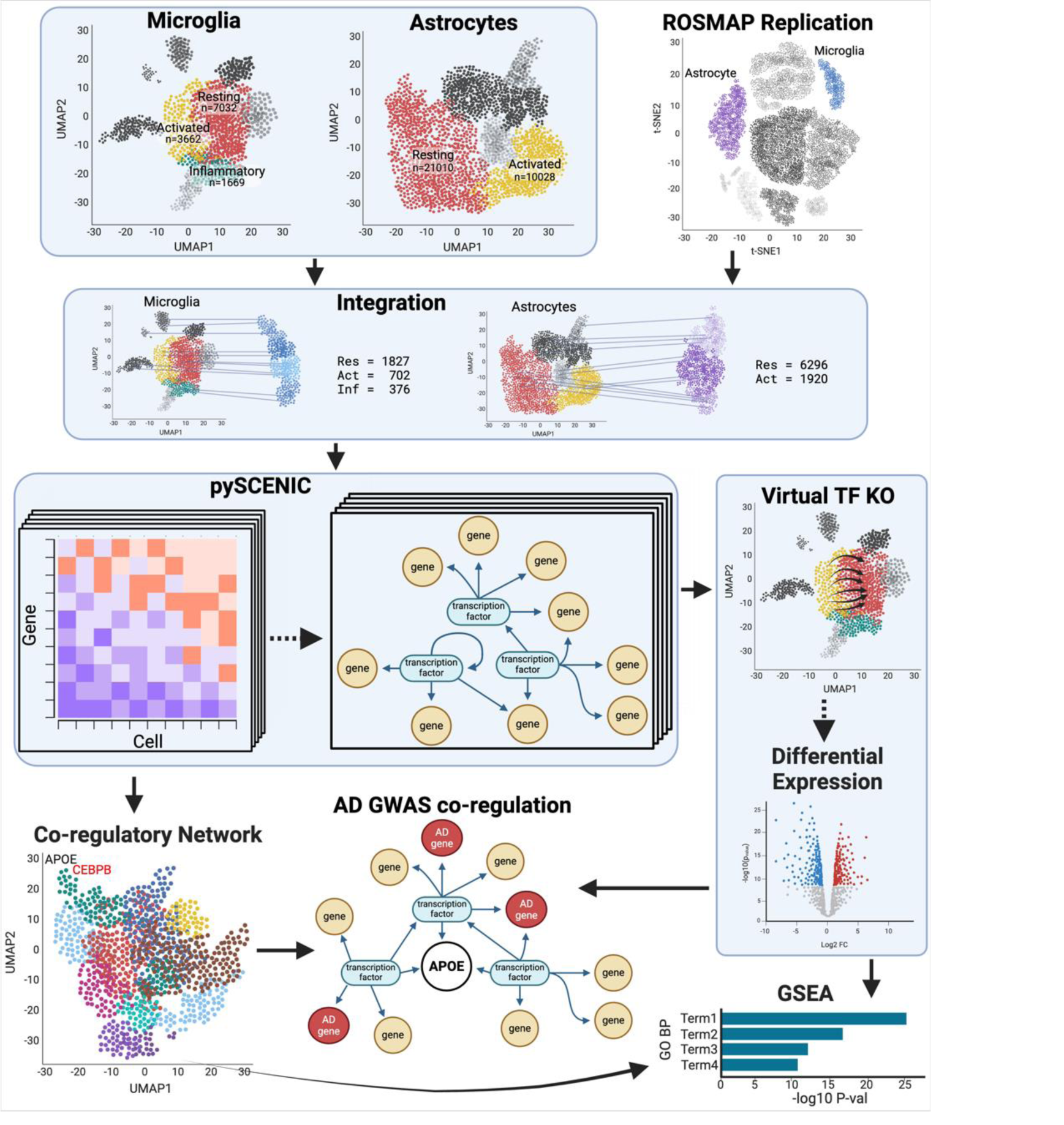
Analysis workflow. We extracted microglial and astrocytic cell states and ran them individually through pySCENIC to create state-specific GRNs. We replicated the putative pySCENIC-TFs by integrating microglia and astrocytes from the ROSMAP cohort and running pySCENIC on them. We clustered the target genes into co-regulatory networks and virtually knocked out the transcription factors (TF KO) that regulated APOE. We identified genes co-regulated with APOE using co-regulatory network clusters and TF-KO DEGs. Both sources of gene sets were tested for inclusion of AD GWAS genes and gene set enrichment.

## Results

### Cell-state-specific transcription factors modulating APOE expression

To identify the state-specific TF regulators of *APOE*, we utilized snRNA-seq data from the Knight ADRC and DIAN cohorts (parietal, n=67)^5^, and replicated the TFs in snRNA-seq from the ROSMAP cohort (dorsolateral prefrontal cortex, n=32)^6^. Focusing on microglia and astrocytes, the principal *APOE* expressors^1^, we isolated the resting and activated states for each cell type: mic-resting (n=7,032), mic-activated (n=3,662), mic-inflammatory (n=1,669), astro-resting (n=21,010), and astro-activated (n=10,028). We leveraged pySCENIC^7^ to build a GRN for each state, and detected an average of 117 and 109 reproducible TF regulators for Knight and ROSMAP cohorts respectively (Supplementary Table 1). We compared the putative TFs between the cohorts and identified an average of 96 replicating regulators for each cell state (Figure 2, Supplementary Table 1). Among these, 35 TFs were shared by all five states, including *ARID3A* (*ABCA7* locus), *GABPA* (*APP* locus), and *MAF* (*WWOX* locus), which are within AD GWAS loci^8^ (Supplementary Table 2). Moreover, 59 TFs were shared by the microglia states, including *SPI1*, *IKZF1*, *TCF3* (*KLF16* locus), and *GABPB1* (*SPPL2A* locus), which are within AD GWAS loci^8,9^, and 69 TFs were shared by the astrocyte states including *SREBF1* (*MYO15A* locus) and *ZNF518A* (*BLNK* locus).

**Figure 2.**
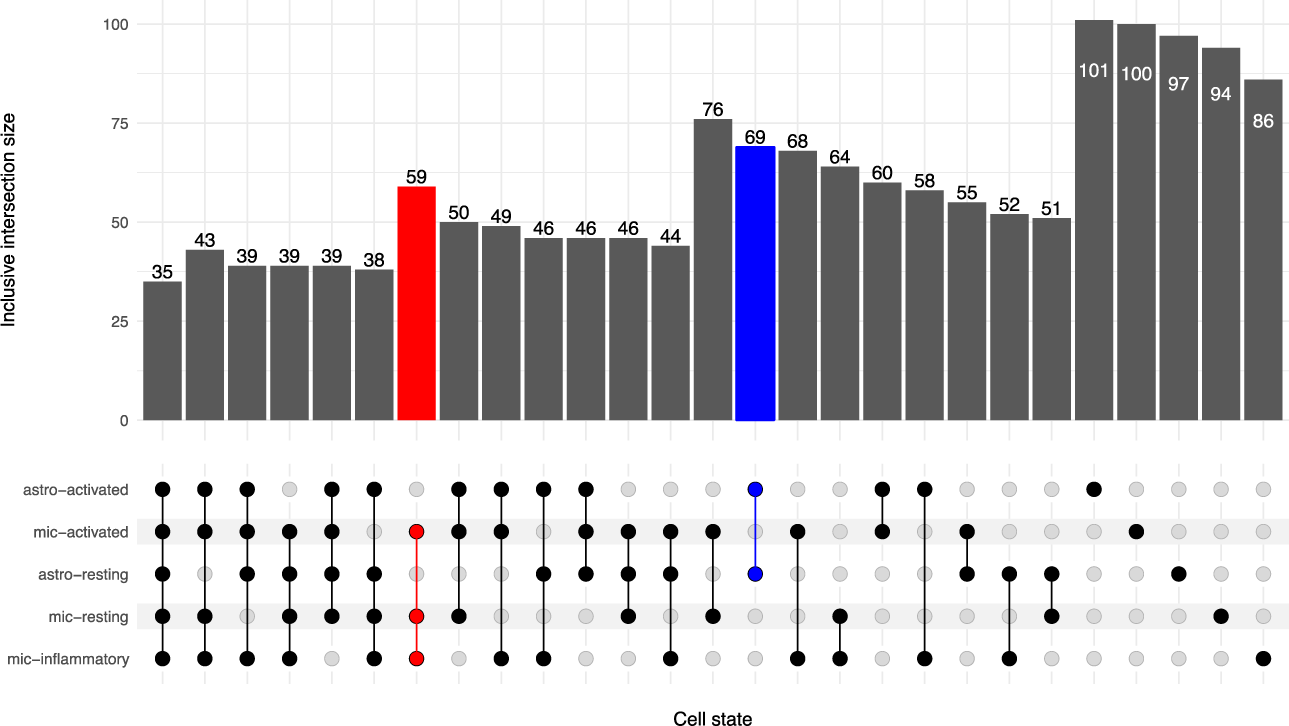
Inclusive intersection of state-specific TF regulators. TF sets were identified by individually running pySCENIC on the Knight/DIAN and ROSMAP cell states and isolating the intersecting TFs between the discovery and replication runs (see Methods). The TF sets were then compared in an upset plot. The inclusive intersection indicates that all subsets are counted in the superset. For example, all 35 TFs shared between all cell states (the lowest subset) are counted in all other sets. The red column indicates the TFs that are common between all microglia states. The blue column indicates that TFs that are common between both astrocyte cell states.

### Different TF families regulate APOE between microglia and astrocytes

Next, we pinpointed the TFs regulating *APOE* in each state. In the mic-resting GRN, APOE was regulated by *CEBPA*, *CEBPB*, *CEBPD*, *IRF7*, and *JUND* TFs. In the mic-activated and mic-inflammatory GRNs, APOE was regulated by *CEBPB*, *CEBPD*, and *JUNB*, with *IRF7* also involved in the mic-activated state. In the astrocytic GRNs, *APOE* was regulated by *JUN* and *THRA* in the resting state and by *THRB* in the activated state.

These results highlight the CCAAT/enhancer-binding protein (CEBP) family of TFs, which form homo- and heterodimers to precisely control the expression of target genes that are primarily immune-related^10^. In mouse microglia cell cultures, *CEBPA* was shown to regulate homeostatic and anti-inflammatory DAM genes^11^, and in mouse models, *CEBPB* modulates *APOE* expression^12^. These findings validate pySCENIC’s ability to capture known biology and lends credence to the other findings. In addition, *CEBPD* was shown to boost the expression of activation genes, including *NOS2*, *C6*, *IL1B*, and *IL6* in a BV2 microglia cell line^13^. Intriguingly, LPS treatment upregulates *CEBPB* and *CEBPD* while downregulating *CEBPA*, implying contrasting regulatory functions. This could account for *CEBPA*’s role as an *APOE* regulator only in mic-resting, but not in mic-activated and mic-inflammatory.

IRF plays a crucial role in the immune response. *IRF7* regulates type I interferon genes and *IL6*^14^. The JUN family proteins are known to be involved in various cellular processes, including proliferation, apoptosis, and immune response^15^. Their role in regulating *APOE* expression suggests a potential link between *APOE* and these cellular processes.

Thyroid hormone receptor family proteins, including *THRA* and *THRB*, bind to thyroid hormone, which plays a crucial role in metabolism, growth, and development. Using cell lines, *THRB* was shown to upregulate *APOE* expression in astrocytes by forming a complex with *RXRA* and binding to the multienhancer ME.2 which activates the *APOE* promoter^16^. Once again, this shows that pySCENIC identifies true regulatory relationships.

Our findings suggest that *CEBPB* and *THRB* regulate *APOE* expression in the human brain, in concordance with studies in model systems. The GRNs from pySCENIC suggest that the CEBP family primarily regulates *APOE* in microglia, while the THR family chiefly regulates expression in astrocytes. This is in line with CEPB regulating immunity genes and microglia being the primary immune cells in the brain, as well as THR influencing metabolism and astrocytes being the primary support cells for neurons. This does not exclude the possibility that both families may influence *APOE* expression in both cell types.

### APOE-associated target genes were regulated by TFs linked to the circadian rhythm

We aimed to explore those genes co-regulated with *APOE*. Thus, we built a complex co-regulatory network of genes, including all the putative TFs predicted by pySCENIC and their respective regulated genes. Then, we clustered the target genes into a UMAP space using the 86-101 TFs signatures that defined each cell state. We identified an average of 27 gene clusters in each expression state (Figure 3, Supplementary Table 1). The gene clusters containing *APOE* comprised an average of 327 genes regulated by an average of 13 TFs for each cell state (Supplementary Table 1).

**Figure 3.**
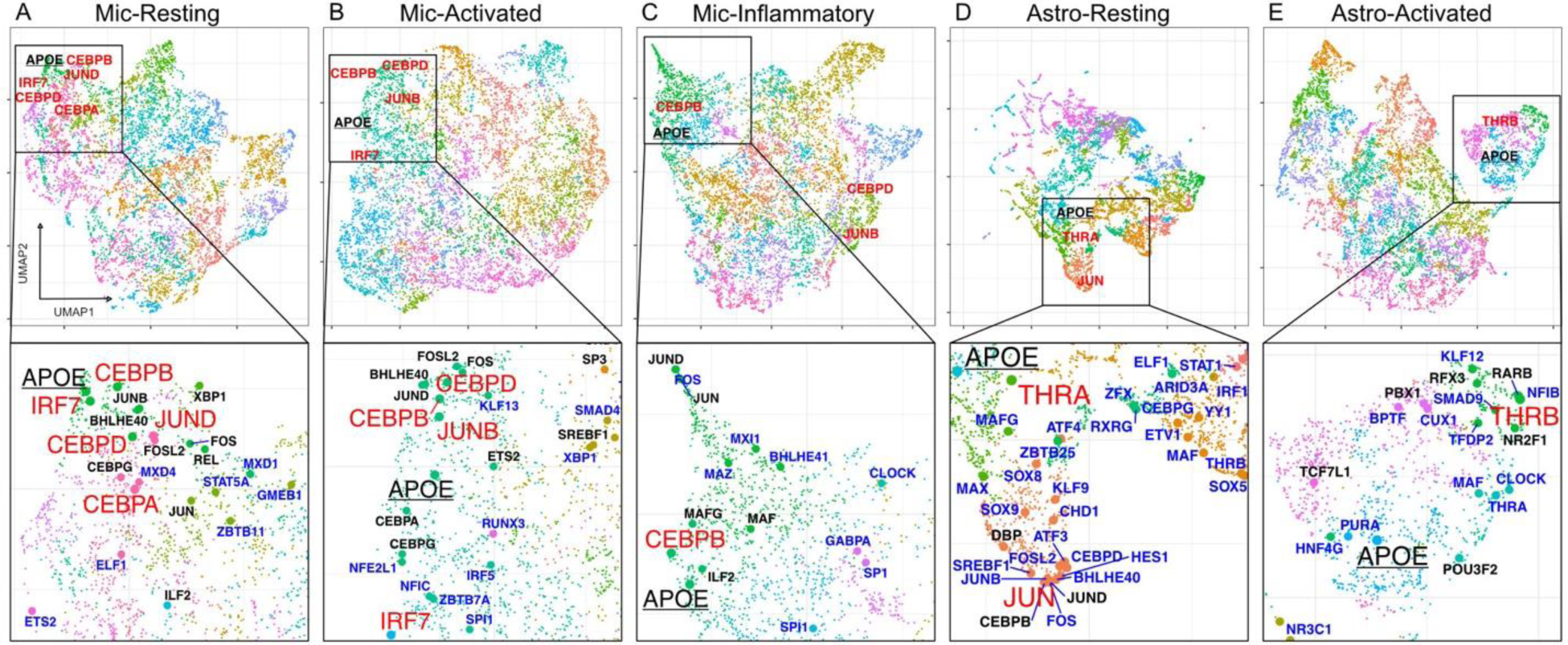
UMAP representation of gene regulatory networks. A-E) A global UMAP visualization of each cell states’ regulome along with a zoomed view highlighting APOE. Principle component analysis was performed on the TF-by-target gene matrix. The top five principal components were used to calculate the UMAP coordinates. APOE is underlined. The TFs in red directly regulate APOE. TFs in black significantly upregulate the APOE gene cluster. TFs in blue are the TFs that were replicated in the ROSMAP dataset. D) The global UMAP for Astro-resting is zoomed. The full figure can be found in Supplementary Figure 3.

To validate these gene clusters, we isolated the TFs that significantly upregulated the gene clusters containing *APOE*, a known activated marker gene, and *CX3CR1* and *P2RY12*, known homeostatic marker genes, in the three microglia clusters. We then performed a virtual knockout (KO) of these TFs and observed the shift in the expression profile of the cells. We adapted the virtual TF knockout method described in SCENIC+^17^ to work with pySCENIC outputs. Following the knockout of the TF, this method propagates the change through the rest of the gene expression matrix (GEM) producing a perturbed expression profile and an associated visualization of the shift in the reduced dimensionality space. The TFs regulating these homeostatic and activated marker genes should have opposite influences on microglial activation, thus opposing shifts in the expression profiles. In the mic-resting state, an individual knockout of the TFs associated with the *APOE* gene cluster generally shifted cells toward the mic-resting state, indicating these TFs are involved in maintaining an activated state. Conversely, individual knockouts of seven of the 14 TFs associated with the *CX3CR1* and *P2RY12* clusters generally pointed away from the mic-resting state, suggesting these TFs promote a resting state (Supplementary Figure 1). A similar trend was seen within mic-activated, with KO of all 17 *APOE* TFs pointing away from mic-activated and eight of the 19 TFs*^CX3CR^*^1/*P2RY12+*^ pointing toward mic-activated (Supplementary Figure 2). This indicates that the clustering groups mirror the expected biology. The mic-inflammatory cluster had no TFs significantly upregulating the *CX3CR1* or *P2RY12* clusters, so it was not explored for validation.

We then explored the TFs regulating *APOE* in each state. The mic-resting *APOE* cluster was regulated by *BHLHE40*, *CEBPG*, *FOS*, *FOSL2*, *GABPA*, *ILF2*, *JUN*, *JUNB*, *KMT2B*, *REL*, and *XBP1*, in addition to the TFs directly regulating *APOE* mentioned previously (Figure 3A, Supplementary Figure 4A). *BHLHE40* regulates the circadian rhythm, negatively regulates itself, *DBP* (another circadian gene), and represses RXR, a component of *APOE* activation through the THR family of TFs^18–21^. BHLHE40 is located within an AD African-American GWAS locus is a key regulator for clearing lipid-rich cellular debris by lipid-associated macrophages (LAMs)^8,22^. *XBP1* becomes a functional TF in response to endoplasmic reticulum stress induced by unfolded proteins and is modulated by the circadian rhythm^23,24^. The mic-activated *APOE* cluster included *BHLHE40*, *CEBPA*, *CEBPG*, *CHD1*, *CREM*, *ETS2*, *FOS*, *FOSL2*, *FOXO1*, *FOXO3*, *JUND*, *SP3*, *SREBF1*, and *USF2*, but not *IRF7*, compared to *APOE* alone (Figure 3B, Supplementary Figure 4B). *CREM* and *SREBF1* are related to circadian rhythm and *SREBF1* is additionally related to lipid metabolism^25,26^. Upregulation of *FOXO3* and two other TFs, was shown to move weakly inflammatory microglia to a strongly inflammatory state in a cell line^27^. Additionally, *FOXO3* is deactivated by *SIRT1*, another gene associated with circadian rhythm^28^. The mic-inflammatory *APOE* gene cluster added *CEBPA*, *E2F4*, *FOSL2*, *GABPB1*, *HDX*, *ILF2*, *IRF7*, *JUN*, *JUND*, *KMT2B*, *MAF*, *MAFG*, *NFE2L1*, *NR3C1*, *TCF7L2*, and *ZBTB40* to the *APOE* TF list (Figure 3C, Supplementary Figure 4C). *E2F4* halts the cell cycle and has been linked to cell quiescence, regulation of AD-related gene networks, and upregulation in AD brains^29^. Macrophage/microglia activation factor (*MAF*) expression has been linked to changes in the endosome/lysosome membranes^30^, which suggests a concerted action between *APOE* and endo-/lysosomal function in activated microglia. Taken together, the CEBP, FOS, and JUN families regulate the *APOE* gene clusters across microglial cell states.

The *APOE* clusters within astrocytes were regulated by fewer TFs than their microglial counterparts. Interestingly, astro-resting did not include either of the THR-related TFs. However it included another steroid/thyroid hormone family receptor *NR2F1*^31^ (Figure 3D, Supplementary Figure 4D), which is associated with Bosh-Boonstra-Schaaf optic atrophy (BBSOA), optic nerve atrophy featured by the loss of retinal ganglion cells critical for the light entrainment of the circadian rhythm^32,33^. The *APOE* cluster in astro-resting also had TFs *DBP*, *CEBPB*, *ING4*, *JUND*, and *TBP* (Figure 3D, Supplementary Figure 4D). This supports the idea that the CEBP family of TFs influences *APOE* in astrocytes and microglia. As previously noted, *DBP* is related to the circadian rhythm. Astro-activated had *MAFG*, *NFIC*, *NR2F1*, *PBX1*, *POU3F2*, *RARB*, *RFX3*, and *TCF7L1* regulating the *APOE* gene cluster in addition to *THRB* which directly regulated *APOE* in this cell state (Figure 3E, Supplementary Figure 4E). *MAFG* has been implicated in astrocyte-driven inflammation linked to the chronic inflammatory disease, multiple sclerosis^34^. *TCF7L1* is important for astrocyte maturation, highlighting the central role that *APOE* plays in normal astrocyte function^35^. *RARB* is another steroid/thyroid hormone family receptor that forms heterodimers with RXR to facilitate transcriptional activation or repression. Despite the absence of the THR-related TFs in astro-resting, both *APOE* gene clusters were regulated by steroid/thyroid hormone receptors, suggesting expression regulation through thyroid hormone. Overall, we observed that circadian rhythm-related TFs were associated with regulating the *APOE* gene clusters across cell types and states. This implies that *APOE* participates in processes similar to those regulated by these TFs, even if they do not directly regulate *APOE*. Alternatively, these TFs may have a role in APOE regulation, but the pySCENIC analysis may lack the power to detect them.

### Gene set enrichment of APOE gene clusters

To investigate the pathways behind the genes within the *APOE* gene clusters from the co-regulatory networks, we performed an enrichment analysis using Gene Ontology (GO) biological process (BP) terms. Mic-resting showed an enrichment of pathways related to cytoplasmic translation, cytokine production, apoptotic process, integrated stress response, immunoglobulin-mediated immune response, response to lipid, IL-6 production, and response to LPS (Supplementary Figure 4F, Supplementary Table 3a). Mic-activated shared most of these pathways, but also the glycolytic process (Supplementary Figure 4F, Supplementary Table 3b), which is a metabolic feature of AD^36^. Mic-inflammatory had fewer pathways than the other two microglial states, but they included endocytosis, macroautophagy, and cotranslational protein targeting to the membrane (Supplementary Figure 4F, Supplementary Table 3c). These results imply that the genes in this cluster are related to the uptake and degradation of extracellular material and the replenishment of membrane proteins.

Astro-resting was enriched for genes in mitochondrial electron transport, NADH to ubiquinone and other aspects of the electron transport chain. It also had genes related to Golgi vesicle transport and synapse organization (Supplementary Figure 4F, Supplementary Table 3d). These results reflect the roles of astrocytes in energy metabolism and synaptic modulation. Astro-activated had many genes in its *APOE* cluster (n=409), which made the enrichment analysis less sensitive. None of the GO terms passed multiple testing correction, but the top hits (p-value < 0.01) included terms such as semaphorin-plexin signaling, axon guidance, synapse assembly, fatty-acyl-CoA biosynthetic process, glucan catabolic process, and amide metabolic process (Supplementary Table 3e). Semaphorin-plexin signaling, in conjunction with axon guidance and synapse assembly, suggests an active regulation of the surrounding synapses. Glucan catabolism indicates the cell is low on glucose and tapping into glycogen reserves. Fatty-acyl-CoA biosynthesis and amide metabolism both indicate that resources are being diverted from ATP production through cellular respiration toward the synthesis of lipids like phospholipids and cholesterol for membranes and neurotransmitters, which is consistent with previous findings in AD^37^.

Overall, all microglia states were associated with immune, inflammation, and cytokine-related terms. We observed that mic-activated and astro-activated cells had genes related to known metabolic alterations in AD. We also noticed that mic-inflammatory and astro-activated cells had genes involved in endocytosis and autophagy, which are mechanisms for clearing amyloid - beta plaques and recycling other macromolecules.

### Gene set enrichment of TF target genes

To complement the analysis of the *APOE* gene clusters, we took a deeper dive into the TFs directly regulating APOE. Specifically, we identified the target genes significantly perturbed by a virtual knockout of these TFs. As previously described, we implemented a modified version of the TF-KO method included in SCENIC+, producing a perturbed expression profile after TF-KO. We then performed a Mann-Whitney rank-sum analysis to identify differentially expressed genes between the original and TF-KO GEMs. We found 60 to 434 nominal DEGs for each TF knockout (Supplementary Table 1, Supplementary Table 4). Using the nominal DEGs, we performed a gene set enrichment analysis and visualized the results in a heatmap to better identify patterns in the regulation of biological processes.

*CEBPB*-regulated processes were common across the microglial states, including positive regulation of cytokine production, receptor-mediated endocytosis, and humoral immune response mediated by circulating immunoglobulin (Figure 4, Supplementary Table 5). These processes reflect the role of *CEBPB* in modulating the inflammatory response of microglia. However, *CEBPB* also regulated processes that were specific to certain microglial states. For example, in mic-inflammatory, *CEBPB*-regulated genes are involved in mRNA splicing, via spliceosome and cellular response to lipopolysaccharide (Figure 4, Supplementary Table 5). In contrast, in mic-activated, *CEBPB* lacked the terms related to positive regulation of protein phosphorylation, positive regulation of transferase activity, antigen processing and presentation of exogenous peptide antigen, and cellular response to cytokine stimulus (Figure 4, Supplementary Table 5).

**Figure 4.**
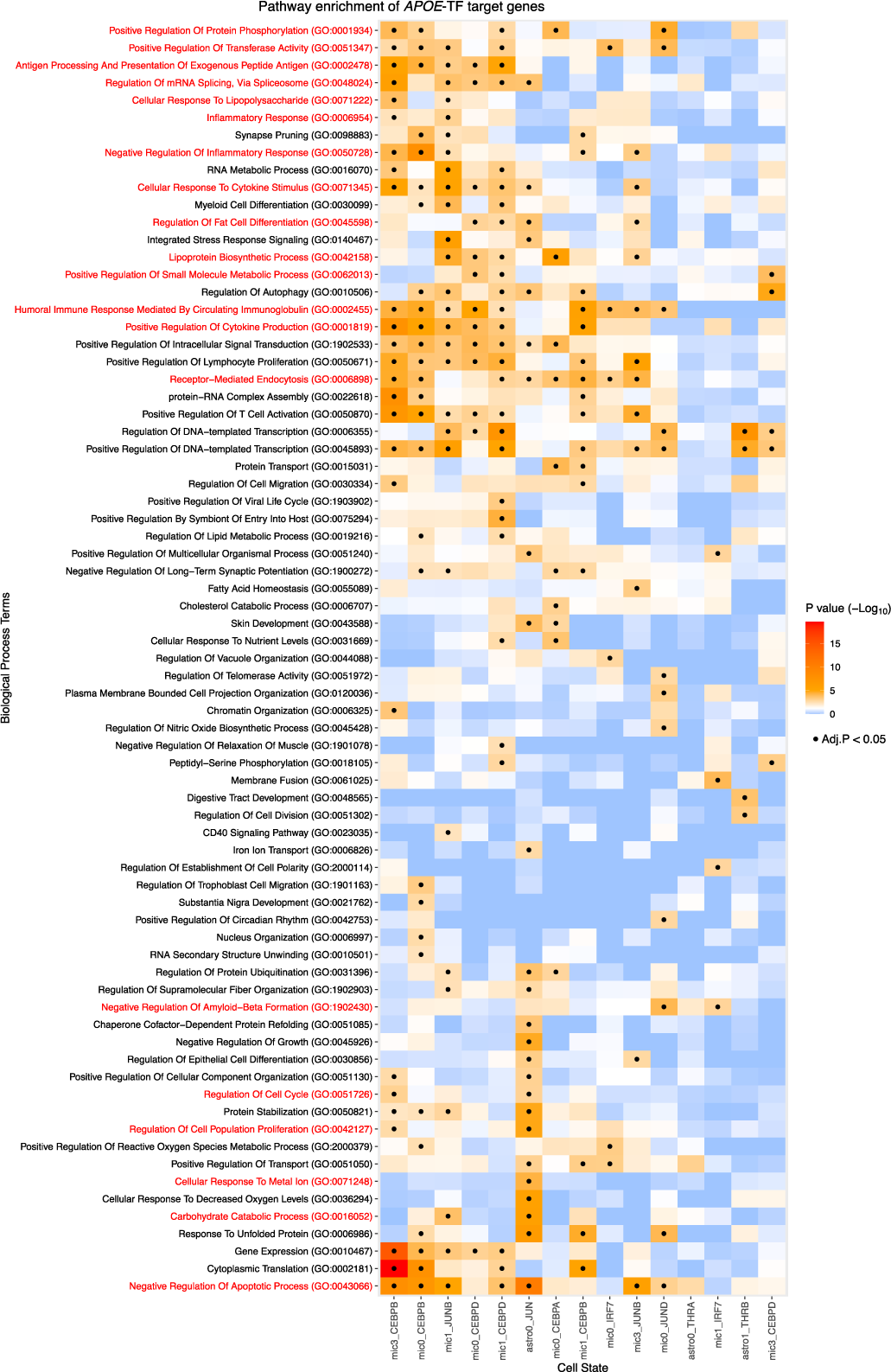
Gene set enrichment analysis of the target genes for each cell-state-specific TF regulating APOE. Target genes for each TF were assessed for GO Biological Process terms. Terms were condensed using Revigo. Rows and columns were sorted using hierarchical clustering on Euclidean distance. Tiles with (.) indicate significance after multiple testing correction.

*CEBPD* regulated shared processes in mic-resting and -activated, but had a distinct signature in mic-inflammatory. The main divergence between the primary *CEBPB* and *CEBPD* signatures was that *CEBPD* additionally regulated fat differentiation, lipoprotein biosynthesis process, and positive regulation of small molecule metabolic process (Figure 4, Supplementary Table 5). Mic-activated *CEBPD* also regulates the lipid metabolic process. These results suggest that *CEBPD* has a more specialized role in regulating genes related to lipid processing than *CEBPB*.

*JUNB* regulated processes related to the immune response in mic-resting and mic-activated states, including negative regulation of the apoptotic process, negative regulation of inflammatory response, cellular response to cytokine stimulus, and humoral immune response mediated by circulating immunoglobulin (Figure 4, Supplementary Table 5). Within mic-activated, it regulated inflammatory response, cellular response to lipopolysaccharide, but also synapse pruning and regulation of mRNA splicing, via splicesome indicating that *JUNB* regulates additional processes other than immune roles in microglia. *JUND* was the regulator in mic-resting state that also regulated negative regulation of apoptotic process and humoral immune response mediated by circulating immunoglobulin, but uniquely among the JUN family, it also regulated negative regulation of amyloid-beta formation through *RIN1* and *RIN3*, which negatively regulate *BACE1*. This implies that *JUND* may be beneficial in preventing amyloid-beta accumulation in AD. Within astro-resting, *JUN* was the regulator and it had the most divergent profile, regulating processes involved in negative regulation of growth, regulation of cell cycle, regulation of cell population proliferation, cellular response to metal ion, and carbohydrate catabolic process (Figure 4, Supplementary Table 5).

*THRA* target genes were not enriched for any particular GO term, but *THRB*-regulated genes associated with transcription, negative regulation of inclusion body assembly, positive regulation of neurogenesis, cellular response to oxidative stress, axonogenesis, and regulation of cytokinesis (Figure 4, Supplementary Table 5). These processes suggest that *THRB* regulates genes that prepare astrocytes for activation and inflammatory response. Overall, this alternative method of examining the genes co-regulated with *APOE* revealed similar immune, lipid, and energy metabolism pathways to those of the *APOE* gene clusters, strengthening the validity of these biological pathways with *APOE*.

### Additional AD GWAS genes are co-regulated with *APOE*

The identified TFs that regulate *APOE* also regulate other genes in AD GWAS loci. Using the same set of nominal DEGs identified after the TF KO, we isolated the DEGs that intersected the list of genes within AD GWAS loci. Several of the genes are regulated by multiple *APOE* TFs including *PGRN*, *FCGR3A*, *CTSH*, *ABCA1*, *MARCKS*, *CTSB*, *SQSTM1*, *TSC22D4*, *FCER1G*, and several of the HLA genes (Figure 5A,B, Supplementary Figure 5).

**Figure 5.**
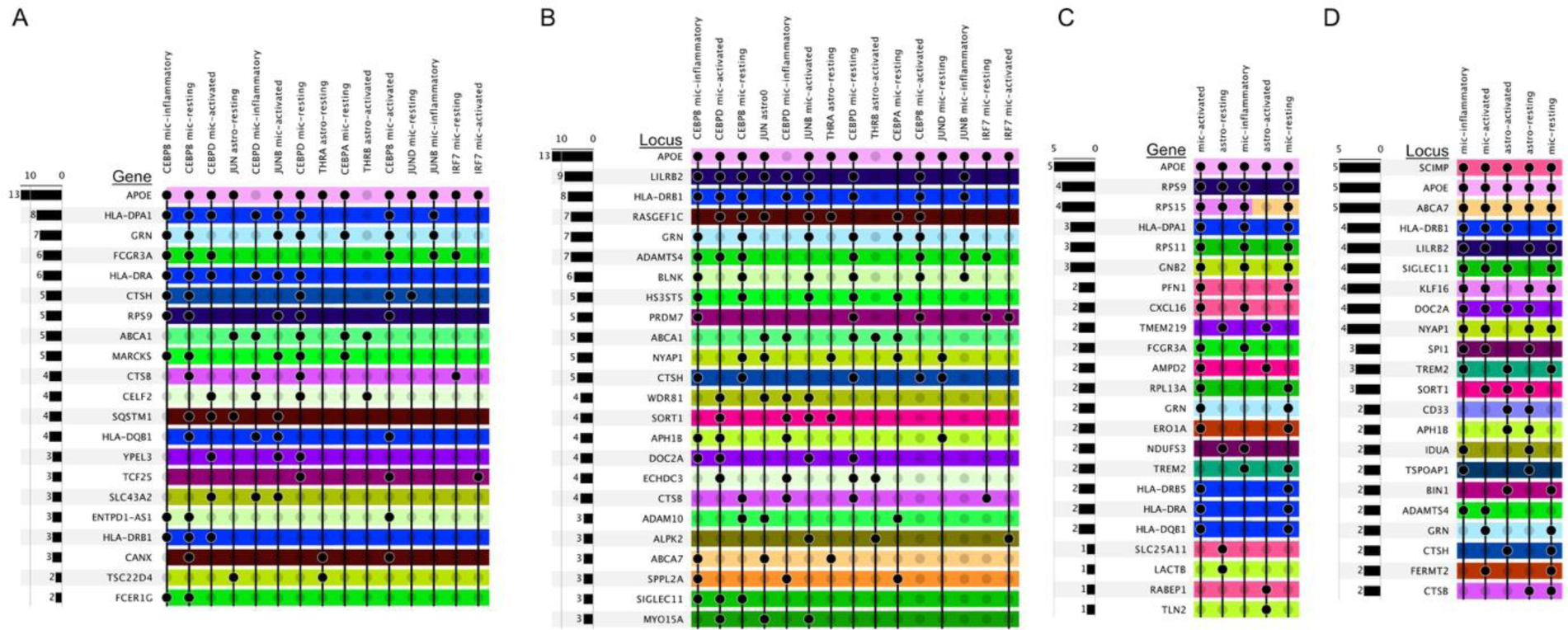
Other AD GWAS genes are coregulated with APOE. (A,B) Genes (A) in AD GWAS loci (B) that are co-regulated by APOE-regulating TFs. Genes co-regulated by APOE-TFs were those differentially expressed (Mann-Whitney Rank U) between the original expression profiles and the perturbed values following virtual TF-KO. (C,D) Genes (C) in AD GWAS loci (D) that co-clustered with APOE in the co-regulation networks. Principle component analysis was performed on the TF-by-target gene matrix. The top five principal components were used to calculate the UMAP coordinates. Gene clusters were calculated from a shared-nearest-neighbor graph using the Louvain algorithm. Rows are colored by loci. Some genes are within two loci and therefore have two colors. These plots are truncated; the full plots can be found in Supplementary Figure 6.

*PGRN, CTSH,* and *CTSB* are all related to lysosome function and are co-regulated with *APOE* in the microglia states. *CTSB* is specifically associated with the proteolytic processing of *APP*^38^. *FCER1G* and *FCGR3A* are in the *ADAMTS4* locus and encode Fc immunoglobulin receptors that bind to IgE and IgG respectively. *CEBPB* regulates *FCER1G* in mic-resting and mic-inflammatory. It represses AD risk genes through the Herpes simplex (HSV-1) escape strategy^39^. *FCGR3A* exacerbates neurodegeneration and is a potential link behind the increased AD risk in chronic periodontitis patients^40,41^. It is regulated in mic-resting by *CEBPB* and *IRF7*; mic-activated by *CEBPB* and *CEBPD*; and mic-inflammatory by *CEBPB*, and *JUNB*. *ABCA1* is a transporter that transfers cholesterol and other lipids to *APOE* and there is a link between *ABCA1*, *APOE*, and Aβ levels^42^. *APOE* and *ABCA1* are co-regulated in both astrocyte and microglial states. *MARCKS* is associated with lipid rafts, dissociates from the membrane at decreased concentrations of cholesterol, causing PIP2 release, and also influences inflammation^43–45^. *SQSTM1* (p62), found in the *RASGEF1C* locus, is also found in both astrocytes and microglia and functions as a bridge between ubiquitinated proteins and autophagosomes^46^. *TSC22D4* is within the *NYAP1* locus and is regulated in astro-resting by both *APOE* TFs (*THRA* and *JUN*). *TSC22D4* participates in forming a complex that degrades *BRI2* and *BRI3*, inhibitors of Aβ production and aggregation.

Looking at the *APOE* gene clusters, we identified genes within the *ABCA7* and *SCIMP* loci across all five cell states and four out of the five states had *HLA-DRB1*, *LILRB2, SIGLEC11*, *KLF16*, *DOC2A*, and *NYAP1* (Figure 5D, Supplementary Figure 5, Supplementary Figure 6). Using *SCIMP* as an example, mic-resting had gene *PFN1* within the APOE gene cluster; mic-activated had *CXCL16* and *PFN1*; mic-inflammatory had *CXCL16*; astro-resting had *SLC25A11*; and astro-activated had *RABEP1* (Figure 5C). Interestingly, *PFN1* is related to amyotrophic lateral sclerosis (ALS), action polymerization, and binding of PIP2^47,48^, which could connect it to *MARCKS* and, therefore membrane cholesterol concentrations as mentioned above. *CXCL16* induces a chemokine-induced chemotactic response, *SLC25A11* maintains the organization and morphology of the mitochondrial cristae^49^, and *RABEP1* is involved in endocytic membrane fusion and recycling of endosomes, all of which connect to AD biology. Genes in AD loci *CD33* and *APH1B* were exclusively found in both astrocyte states (Figure 5D). *LACTB* (APH1B locus) was co-regulated with APOE in astro-resting (Figure 5C) and regulated mitochondrial lipid metabolism^50^. *TLN2* (*APH1B* locus) was co-regulated in astro-activated (Figure 5C) and is a component of the actin cytoskeleton linking to focal adhesion plaques, which have been shown to have higher concentrations of cholesterol than the surrounding membrane^51^. Even though the genes were not necessarily the same across the cell states, the consistency of genes within these loci clustering with *APOE,* and many relating to lipid metabolism, suggests common regulatory mechanisms behind these loci and *APOE*.

These findings suggest that the regulatory networks involving *APOE* and the identified transcription factors extend beyond *APOE* itself and involve other genes associated with AD. Many of these AD-GWAS genes are related to critical immune, lysosome, lipid, and energy metabolism pathways highlighted by the previous TF and gene set enrichment analyses. It also supports the notion that some GWAS loci have many genes that can simultaneously influence AD risk.

## Discussion

We used GRNs to elucidate the cell-type- and -state-specific regulation of *APOE*. These analyses revealed that *APOE* and its co-regulated genes were modulated by several transcription factor (TF) families, including CEBP, JUN, FOS, FOXO, and THR. The CEBP (A/B/D/B), JUN (/B/D), and FOS (/L2) families were involved in *APOE* regulation in all microglial states, whereas FOXO (1/3) was specific to the mic-inflammatory state. Despite the consistency of the CEBP, JUN, and FOS families across microglial states, the individual members had different strengths and associations based on cell state suggesting individual TFs could more strongly promote one state over another. The CEBP family of TFs form homo- and heterodimers with each other, so the proportions of these dimers could influence cell state. Additional experimental studies are needed to better understand the interplay between members of these families and how they regulate the activation states of microglia. Interestingly, *CEBPB* and *JUND* also regulated the *APOE* gene cluster in the astro-resting state, suggesting that these TF families play a role in *APOE* regulation in both microglia and astrocytes. *APOE* in stro-activated was controlled by the known *APOE* regulator *THRB*^16^, and *NR2F1* and *RARB*, two other members of the steroid/thyroid hormone receptor family. We previously reported an upregulation of *THRB* in the mic-activated state compared to the other microglia states, implying that the THR family of TFs could influence *APOE* expression in microglia as well, but to a lower degree^5^. Further studies are required to confirm this conclusion.

These GRNs identified associations with immunoglobulin binding, IL1B/IL6/cytokine production, glycolysis, endocytosis, macroautophagy, mitochondrial electron transport, fatty-acyl-CoA biosynthesis, glucan catabolism, and amide metabolism. Related to immunoglobulin binding, AD GWAS associated genes *FCER1G*, *FCGR2A*, and *FCGR3A* are Fc immunoglobulin receptors that bind to IgE and IgG and were coregulated with *APOE* in microglia. Usually, IgE and IgG cannot cross the blood-brain-barrier (BBB), but it has been suggested that in *APOE*e4 carriers, the vasculature and endothelial cells making up the BBB are compromised, and albumin, antibodies, cytokines, and other inflammatory molecules more easily cross into the brain^52,53^. This might account for the association between this biology and *APOE*. It might also explain how periodontitis, a peripheral infection, could increase AD risk in association with *FCGR3A*^41^.

Additionally, when microglia and astrocytes encounter these inflammatory signals, they propagate the signal by producing their own *IL1B*, *IL6*, and other cytokines, as suggested by these GRNs^54–56^. Astrocytes also boost their cholesterol production in response to cytokine signaling^53^. Within astro-activated, we saw many *APOE*-related genes pointing to fatty-acyl-CoA biosynthesis, a precursor to cholesterol. *APOE* is one of the key molecules transporting this newly synthesized cholesterol to the surrounding cells. Evidence indicates that increases in cholesteryl ester, the storage molecule for excess cholesterol, increase the accumulation of phosphorylated tau (pTau) by reducing the proteasome protein units^57,58^. Studies have also suggested that this increased cholesterol enlarges the lipid rafts, influencing many associated receptors^53,59,60^. For instance, *APP*, *BACE1*, *TREM2*, *MS4A*, *TLR4*, Fc immunoglobulin receptors, and *GLUT1* are all associated with lipid rafts^53,59–61^. Our analysis shows that other AD GWAS genes are co-regulated with APOE; some are also associated with cholesterol levels in the membrane, including *MARCKS*, *PRN1*, and *TLN2*. In neurons, *APP* is upregulated and preferentially beta-cleaved when associated with lipid rafts which is amyloidogenic^59,60^. *TREM2*, *TLR4*, *IFNGR*, and *TNFA* are all receptors that rely upon clustering as a part of their activation^53^, and increased cholesterol mediates this clustering, enhancing the activation of these cells, again highlighting the connection between cytokines and the *APOE*-related genes. Astrocytes are the primary support cells of the brain and are in contact with the vasculature to facilitate the uptake and delivery of essential molecules to the neurons and other cells. *GLUT1* transports glucose into astrocytes through the endfeet, but evidence suggests that *GLUT1* is less efficient when incorporated into large lipid rafts induced by increased cholesterol^62,63^. This might explain why the mitochondrial electron transport chain is associate with astro-resting, while glucan catabolism, or the breakdown of glycogen into glucose, is linked to astro-activated state. Other studies have reported a reduction in glucose uptake in AD^36^. Interestingly, while extra cholesterol decreases glucose uptake, it increases neuronal energy demand by promoting neurotransmitter release in presynapses and the endocytosis and internalization of receptors in postsynapses^64^. This energy demand stimulates the transport of lactate and ketones from astrocytes to neurons, enabling increased neural activation by bypassing the rate-limiting step of glycolysis in energy production^36,65^. Evidence shows that in young *APOE*e4 carriers, there is an increase in cognitive performance and neuronal activity, suggesting this biology is occurring^36,66,67^. With age we see the negative aspects of this increased neuronal activity. Neuronal activity produces reactive oxygen species (ROS), so these hyperactive neurons produce increased quantities of ROS which then peroxidate the fatty acids (pFA) found in the cell and organelle membranes^68^. These pFA are toxic, so the neurons expel them with the help of *APOE*. Over time, ROS and pFA build up in the neuron, causing neurotoxicity, especially in APOEe4 carriers with less efficient binding and, therefore, export of lipids. The expelled pFA is then picked up by microglia and astrocytes and stored in lipid droplets (LD), which is consistent with our finding GO terms, TFs (*MAF* and *CEBPB*), and GWAS genes (*RABEP1*) related to endocytosis. This is also concordant with our finding that *BHLHE40*, a regulator for the clearance of lipid-rich cellular debris, is an *APOE* regulator. Excess cholesterol produced by astrocytes also collects in the lysosome, where it causes lysosomal dysfunction, leading to the buildup of autophagosomes and mitochondrial dysfunction^36^. This is consistent with many of the co-regulated GWAS genes we identified, including *PGRN*, *CTSH*, *CTSB* (lysosome), *SQSTM1* (autophagosomes), *SLC25A11*, and *LACTB* (mitochondria metabolism).

Our findings also show that many TFs regulating *APOE* and the *APOE* gene clusters are related to circadian rhythm, including *BHLHE40*, *DBP*, *XBP1*, *CREM*, *SREBF1*, *FOXO3*, and *NR2F1*. This is consistent with many studies that have reported an association between AD and disrupted sleep due to circadian rhythm dysfunction^33,66,69–71^. These findings highlight the importance of the circadian rhythm in regulating APOE expression and function. Interestingly, *NR2F1* is associated with BBSOA, a neurodevelopmental disease characterized by the loss of retinal ganglion cells^32^. Another study with *APOE*^-/-^ reported the loss of melanopsin-expressing ganglion cells in the suprachiasmic nucleus, resulting in the loss of light entrainment^33^, the process by which light synchronizes the biological clock with the environment. These studies suggest that *NR2F1* and *APOE* are connected and involved in the circadian rhythm, as indicated by our GRNs. Furthermore, the disruption of energy metabolism and the suggested increase in ketone metabolism are modulated by *AMPK*^72^. *AMPK* also degrades the circadian proteins *Per* and *Cry*, which are produced by the transcriptional activation of *Clock* and *Bmal1*^73^. *Per* and *Cry* form a complex with *CK1E/D* and inhibit the transcription of *Clock*, *Bmal1*, *Per*, and *Cry*, creating a negative feedback loop. The degradation of *Per* and *Cry* by *AMPK* breaks this loop and results in the overexpression of *Clock* and *Bmal1*, as well as their downstream targets, such as *NR1D1*, which encodes the Rev-ERBα protein. *NR1D1* represses *Cx3cr1*, a marker of homeostatic microglia, thus activating microglia and influencing lipid droplet formation^74,75^. Disruption of the circadian rhythm is associated with impaired function and integrity of the endothelial and vascular smooth muscle cells, causing inflammation and passing these inflammatory markers through a weakened BBB^73^. It is also related to energy metabolism during the fasting stage of sleep^26^. Overall, our results reemphasize the connection of AD to circadian rhythm and suggest that *APOE* and lipid biology are key contributors.

In general, the TF families primarily associated with microglia suggest a relationship between immunity and *APOE*, while the TF family associated with astrocytes suggests a link between *APOE* and lipid metabolism. This is consistent with the primary function of each of these cell types. In astrocytes, the evidence points to the primary purpose of *APOE* in lipid metabolism is to shuttle cholesterol and other lipids synthesized by the astrocytes to other cells. The primary purpose of *APOE* in immunity is not quite as clear in microglia. A promising possibility is the secretion of extracellular vesicles (EVs) that are induced upon microglial activation. The biology related to these EVs is closely tied with that of the endosomal-lysosomal system^76^, highlighted in our GRNs. These EVs can contain RNA, protein, lipids, and cytokines for extracellular communication^77^. A study in *Apoe*^-/-^ mice on a high-fat diet highlighted the role of *Apoe* in influencing the miRNA composition of EVs secreted by these macrophages and their impact on atherosclerosis^78^. Although most studies on EVs interrogate their miRNA composition, other studies suggest differences in lipid composition based on microglia state as well^79^, including increased secretion of cholesterol in EVs in response to increased free cholesterol^80^. *APOE* could play a role in the transfer of lipids to these EVs.

A limitation of this study is that pySCENIC utilizes only a single modality, co-expression, to identify relationships between TFs and target genes, unlike the newer SCENIC+, which also incorporates chromatin accessibility. Despite this, pySCENIC has successfully identified biologically relevant TF-target interactions. We presume that TFs regulating the *APOE* gene clusters within the co-regulatory networks influence APOE, even in the absence of direct TF-*APOE* links in the pySCENIC results. Nevertheless, the findings underscore many known features of AD and establish a link to *APOE* biology. Moreover, the discovery cohort includes carriers of *APOE*, *TREM2*, *APP*, and *PSEN1* variants, all of which could influence the GRNs in ways not accounted for. Additionally, the cohort’s European ancestry may restrict the generalizability of these results to other, less represented populations.

In conclusion, the TFs and pathways associated with *APOE* all support known AD pathology and support the building evidence implicating lipids as a key driver of AD-related pathology.

## Methods

### Single nucleus RNA-seq acquisition and integration

Human snRNA-seq data of the parietal lobe from the Knight ADRC and DIAN was gathered at NIAGADs (accession number <u>NG00108</u>) and upon request from DIAN. These data include a total of 67 neuropathological controls, sporadic AD, and autosomal dominant AD participants. An independent human snRNA-seq dataset from ROSMAP (dorsolateral prefrontal cortex) was collected from Synapse (synapse ID <u>syn21125841</u>). The ROSMAP data has 11 sporadic AD, 11 *TREM2* R62H, and 10 control participants. The microglia from the ROSMAP dataset was previously integrated with the microglia from the Knight and DIAN data^5^. The ROSMAP astrocytes were integrated as a part of this study using the same process described by Brase, *et al*. Briefly, we isolated and normalized the astrocytes with Seurat’s (v.4.3.0) *SCTransform*, setting “return.only.var.genes” to FALSE and regressing out “nCount_RNA” and “nFeature_RNA”. We integrated the astrocytes with 3000 features in *SelectIntegrationFeatures*, *PrepSCTintegration*, our data as reference in *FindIntegrationAnchors*, and *IntegrateData*. We clustered integrated data with ten principal components in *FindNeighbors* and resolution 15 in *FindClusters*. We gave each cluster an ‘original’ identity by isolating our nuclei from clusters and finding the most common original ID. We transferred this ID to ROSMAP nuclei like a k-nearest neighbor classifier. We mapped cluster identities to pre-integrated normalized ROSMAP data.

### Gene regulatory network prediction by pySCENIC

We created cell-state-specific GRNs using the python implementation of the SCENIC^81^ analysis method called pySCENIC^7^ (version 0.12.1). We isolated the cell type of interest and filtered the genes to those that were expressed in ≥5% of the cell type population. Then we isolated the cell expression states of interest. We ran pySCENIC on each individual cell state 100 times (different seed for each run) as suggested by pySCENIC. We employed default values for all parameters and provided default reference data downloaded from https://resources.aertslab.org/cistarget/: Database (hg38_500bp_up_100bp_down_full_tx_v10_clust.genes_vs_motifs.rankings, hg38_10kbp_up_10kbp_down_full_tx_v10_clust.genes_vs_motifs.rankings), table (motifs-v10nr_clust-nr.hgnc-m0.001-o0.0.tbl), and transcription factor list (allTFs_hg38.txt). An additional TF list was downloaded from http://humantfs.ccbr.utoronto.ca/download/v_1.01/TF_names_v_1.01.txt and the two lists were merged totaling 2093 TFs.

After the 100 runs were complete, we identified the transcription factors (TFs) that were present in >= 80% of the runs. We then ran pySCENIC an additional 100 times using only the >=80% TFs labeled as transcription factors. The other TFs were retained in the expression matrix, but not labelled as TFs. The TFs that appeared in >= 80% of the second round of runs were isolated as reproducible regulators.

The same resource files and parameters were used in the ROSMAP cohort replication. We filtered the genes to those intersecting those from the Knight ADRC microglia and astrocyte runs. We ran pySCENIC 100 times using the filtered list of TFs for each respective cell state (list used in second round of discovery runs). We isolated the TFs that were present in >= 50% of the runs. We intersected the final lists of discovery TFs with the lists of replication TFs to identify a set of replicable TFs for each cell state.

### Co-regulation network clusters

Using the TF-target gene matrices for cell state, we identified co-regulation network clusters and visualized them in the UMAP space. The TF-target gene matrix had each of the replicable TFs as features, the target genes as elements, and the values were the number of times the target gene was regulated by the TF out of the 100 pySCENIC runs. These values were normalized by the number of times the TF was included in the pySCENIC networks. For example, if TF_1 was in the pySCENIC networks 85 times, all the values would be multiplied by (100/85) to get the normalized value. We then log transformed the values and ran them through a principal component analysis using *prcomp,* a native R (v4.2.2) function. We then passed the top five principal components (PCs) through the *umap* function from the umap library (v0.2.10.0) to create a two-dimensional representation of the space. The top five PCs were also passed to the *makeSNNGraph* function from the bluster package (v1.8.0) to create a shared nearest neighbor graph. The resulting graph was passed to the *cluster_louvain* function from the igraph package (v1.3.5) to identify clusters of similarly regulated target genes using a resolution of 2. All target genes in the same cluster as APOE were considered co-regulated with APOE and were evaluated for additional AD GWAS genes and for gene set enrichment (Supplementary Table 6).

### TF regulators for gene clusters

We identified the TFs significantly upregulating the genes in each gene cluster identified in the gene UMAPs. We utilized the normalized, but not log transformed, TF-target gene matrix described in the Gene UMAP clusters section. For each gene cluster at a time, we created a binary vector where 1 indicated the target gene was in the cluster. We then looped through each TF and modelled the TF-target gene values using the binary vector as a covariate. The native *glm* R function (v4.2.2) was used to train the model with a quasipoisson distribution and log link function. The hits for each cluster were then multiple testing corrected using the Benjamini-Hochberg (BH) method. TFs with a BH corrected p-value less than 0.05 and an estimate greater than zero were considered significant regulators for the gene cluster.

### In silico transcription factor knock-out

SCENIC+^17^ was recently released with the functionality to knockout a TF and determine the shift in the expression space. We implemented the algorithm proposed in SCENIC+ (v1.0.0, python v3.8.18), which was an adaption from the original method in CellOracle^82^. In summary, this method uses the cell by gene expression matrix to train prediction models for each gene. These models allow for the virtual knockout of a TF (TF-KO) by setting the expression of the TF to zero and then using the trained models to predict the shift in target gene expression. The expression shift is then transformed back onto the reduction space and depicted by arrows.

By default, SCENIC+ used the GradiantBoostingRegressor from sklearn (v1.3.2) with a learning rate of 0.01, 500 estimators, and max features set to 0.1 to train prediction models. We included the TFs that were regulating a target gene at least 5% of the time in the model that used TF expression to predict the target gene expression. To correct for the influence of sample proportion on each cell state, we also included the log transformed nuclei counts for the sample in the prediction model. SCENIC+ uses a default of five iterations to propagate the shift across the expression matrix. We extended SCENIC+’s implementation to work with the UMAP space rather than just the PCA space. We also switched the arrows to be drawn by the quiver function as was found in the CellOracle implementation rather than the streamplot function used by SCENIC+.

To extend this method to the UMAP space, we trained a neural network to predict a cell’s UMAP coordinates using as input the expression levels of all genes (python v3.11.0). We used the original gene expression matrix (GEM) and UMAP representation for the cell types which included all expression states not just the resting and activated states passed to pySCENIC. First, we log transformed the GEM. We then used the *train_test_split* function from sklearn (v1.3.2) to split the data into training and testing data (80% training, 20% testing) and stratified by cell state. We then split the training data into training and validation sets (90% training, 10% validation) and once again stratified by cell state. Using the *StandardScaler* function from sklearn (v1.3.2) we fit and transformed a scaler using the training GEM. This scaler was then used to transform the validation and test GEMs. The sequential neural network was constructed using tensorflow (v2.14.0) and consisted of three layers: two dense layers with the ReLU activation function and an output layer with the same dimensionality as the UMAP data (n=2). The model was compiled with the Adam optimizer and mean squared error loss function. Early stopping was implemented to prevent overfitting with monitoring set to ‘val_loss’ and a patience of 10. The model was then trained on the training data for a maximum of 1000 epochs. The trained model was evaluated on the test GEM. This trained model was used on the TF-KO perturbed matrices to transform the expression shift to the UMAP reduction space. This method was incorporated into a snakemake pipeline so that TF-KO could be performed consistently across TFs, cell states, and cell types.

### TF knockout differential expression

We performed a differential expression analysis between the original GEM and perturbed GEM calculated after TF-KO. The original GEM and perturbed GEM were both log transformed. We then filtered out the cells that had zero expression of the TF in the original matrix as these cells have no changes in expression and would skew the results. We then performed a Mann-Whitney rank U test using the *mannwhitneyu* function from scipy (v1.2.2, python v3.11.0) to identify differentially expressed genes. We performed FDR multiple testing correction. Genes with a corrected p-value less than 0.05 were considered co-regulated with APOE and were evaluated for additional AD GWAS genes and for gene set enrichment (Supplementary Table 6).

### Gene set enrichment of genes co-regulated with APOE

Sets of co-regulated genes where run through enrichment analysis to better understand the related function. There were two sources of co-regulated genes, co-regulation network clusters and TF-targets, and they were analyzed independently. The gene sets identified by gene clusters were run through enrichment analysis using the *enrichr* function from the enrichR package (v3.1) in R (v4.2.2) and the TF-targets were run through the *enrichr* function from the gseapy package (v0.9.5) in python (v3.11.0). Both used the 2023 GO Biological Process (BP) terms and significant associations were those with adjusted p-values less than 0.05.

The terms from the TF-targets analysis were condensed using rrvgo (v1.6.0), an R package that implements the Revigo^83^ tool for summarizing GO BP terms. The function *calculateSimMatrix* was used to calculate the relationships between the GO BP terms with variable inputs: orgdb = “org.Hs.eg.db”, ont = “BP”, method = “Rel”. The terms were then summarized using *reduceSimMatrix* and the following variables: score = “Rank”, threshold = .7, orgdb = “org.Hs.eg.db”. The summarized terms or “parentTerms” and their P values were then used to make a heatmap. Rows and columns were ordered using dist function method = “manhattan”.

### Identifying other AD GWAS genes co-regulated APOE

There were two sources of co-regulated genes, co-regulation network clusters and TF-targets, and they were analyzed independently. These gene sets were compared against a list of genes within GWAS loci (+/- 500KB of the lead SNP, Supplementary Table 7) and can be found in Supplementary Table 6. The Loci for each source were identified and merged. This merged list was used to consistently color the genes and loci for each row in the upset plots in Figure 5.

## Data Availability

The single nucleus data from the Knight ADRC accessed in this study are found in the National Institute on Aging Genetics of Alzheimer’s Disease Data Storage Site (NIAGADS) with accession number NG00108 [https://www.niagads.org/datasets/ng00108]. The raw single nucleus data from the DIAN brain bank accessed in this study are available under restricted access to maintain individual and family confidentiality. These samples contain rare disease-causing variants that could be used to identify the participating individuals and families. Access can be obtained by request through the online resource request system on the DIAN Website: https://dian.wustl.edu/our-research/for-investigators/dian-observational-study-investigator-resources/. The ROSMAP single nucleus RNA sequencing data used in this study are available at Synapse under Synapse ID syn21125841 [https://www.synapse.org/#!Synapse:syn21125841/wiki/597278]. The pySCENIC default reference data was downloaded from https://resources.aertslab.org/cistarget/: Database [https://resources.aertslab.org/cistarget/databases/homo_sapiens/hg38/refseq_r80/mc_v10_clust/gene_based/hg38_500bp_up_100bp_down_full_tx_v10_clust.genes_vs_motifs.rankings.feather], [https://resources.aertslab.org/cistarget/databases/homo_sapiens/hg38/refseq_r80/mc_v10_clust/gene_based/hg38_10kbp_up_10kbp_down_full_tx_v10_clust.genes_vs_motifs.rankings.feather], table [https://resources.aertslab.org/cistarget/motif2tf/motifs-v10nr_clust-nr.hgnc-m0.001-o0.0.tbl], and transcription factor list [https://resources.aertslab.org/cistarget/tf_lists/allTFs_hg38.txt]. An additional TF list was downloaded from [http://humantfs.ccbr.utoronto.ca/download/v_1.01/TF_names_v_1.01.txt]. Source data are provided with this paper.

## Code Availability

Custom code used to analyze the snRNA-seq data and datasets generated and/or analyzed in the current study are available from the corresponding authors upon request or at https://github.com/HarariLab/APOE-GRN.

## Data Availability

https://dian.wustl.edu/our-research/for-investigators/dian-observational-study-investigator-resources/

https://www.synapse.org/#!Synapse:syn21125841/wiki/597278

https://resources.aertslab.org/cistarget/databases/homo_sapiens/hg38/refseq_r80/mc_v10_clust/gene_based/hg38_500bp_up_100bp_down_full_tx_v10_clust.genes_vs_motifs.rankings.feather

https://resources.aertslab.org/cistarget/databases/homo_sapiens/hg38/refseq_r80/mc_v10_clust/gene_based/hg38_10kbp_up_10kbp_down_full_tx_v10_clust.genes_vs_motifs.rankings.feather

https://resources.aertslab.org/cistarget/motif2tf/motifs-v10nr_clust-nr.hgnc-m0.001-o0.0.tbl

https://resources.aertslab.org/cistarget/tf_lists/allTFs_hg38.txt

http://humantfs.ccbr.utoronto.ca/download/v_1.01/TF_names_v_1.01.txt

## Acknowledgments

Doctors Bruno A. Benitez, and Oscar Harari contributed equally to this work as co-senior authors. Data sharing for this project were supported by The Dominantly Inherited Alzheimer Network (DIAN, U19AG032438), funded by the National Institute on Aging (NIA), the Alzheimer’s Association (SG-20-690363-DIAN), the German Center for Neurodegenerative Diseases (DZNE), Raul Carrea Institute for Neurological Research (FLENI), Partial support by the Research and Development Grants for Dementia from Japan Agency for Medical Research and Development, AMED, and the Korea Health Technology R&D Project through the Korea Health Industry Development Institute (KHIDI), Spanish Institute of Health Carlos III (ISCIII), Canadian Institutes of Health Research (CIHR), Canadian Consortium of Neurodegeneration and Aging, Brain Canada Foundation, and Fonds de Recherche du Québec – Santé. DIAN Study investigators have reviewed this manuscript for scientific content and consistency of data interpretation with previous DIAN Study publications. We acknowledge the altruism of the participants and their families and the contributions of the DIAN research and support staff at each of the participating sites for their contributions to this study.

The results published here are partly based on data obtained from the AD Knowledge Portal (https://adknowledgeportal.org). Study data were provided by the Rush Alzheimer’s Disease Center, Rush University Medical Center, Chicago. Data collection was supported through funding by NIA grants P30AG10161 (ROS), R01AG15819 (ROSMAP; genomics and RNAseq), R01AG17917 (MAP), R01AG30146, R01AG36042 (5hC methylation, ATACseq), RC2AG036547 (H3K9Ac),

R01AG36836 (RNAseq), R01AG48015 (monocyte RNAseq) RF1AG57473 (single nucleus RNAseq), U01AG32984 (genomic and whole exome sequencing), U01AG46152 (ROSMAP AMP-AD, targeted proteomics), U01AG46161(TMT proteomics), U01AG61356 (whole genome sequencing, targeted proteomics, ROSMAP AMP-AD), the Illinois Department of Public Health (ROSMAP), and the Translational Genomics Research Institute (genomic). Additional phenotypic data can be requested at www.radc.rush.edu.

This work was possible thanks to the following governmental grants from the National Institute of Health: NIA R01AG057777 (OH), R56AG067764 (OH), U01AG072464 (OH), R01AG074012 (OH), U19AG032438 (RJB), NINDS R01NS118146 (BAB), R21NS127211 (BAB), NIA T32AG058518 (LB), O.H. is an Archer Foundation Research Scientist.

This work was supported by access to equipment made possible by the Departments of Neurology and Psychiatry at Washington University School of Medicine.

The funders of the study had no role in the collection, analysis, or interpretation of data, in the writing of the report, or in the decision to submit the paper for publication.

Figure 1 was created with BioRender.com. Bing Copilot was used to improve grammar and flow of text.

## Author Contributions

L.B. performed the quality control, bioinformatic analyses, interpreted the results, created the figures, and wrote the manuscript. Y.Y. ran the pySCENIC analyses. E.M. was involved in the DIAN sample collection, profiling and provided a critical review of the manuscript. O.H. and B.A.B. were involved in experiment design, analysis, data interpretation, intellectual contribution, and manuscript writing and review.

## Supplementary Information

Supplementary Information - .pdf file

