## Supplementary Information for "Comparative gene regulatory networks modulating *APOE* expression in microglia and astrocytes"

#### Supplementary Figures

**Supplementary Figure 1 Expression profile shifts following virtual KO of APOE, CX3CR1, and P2RY12 regulating TFs in mic-resting.**
Plots boxed in red indicate those that shift expression away from the resting state. Principle component analysis was performed on the TF-by-target gene matrix. The top five principal components were used to calculate the UMAP coordinates. Gene clusters were calculated from a shared-nearest-neighbor graph using the Louvain algorithm. Linear models were used to identify TFs that significantly regulated the gene clusters. TFs for the clusters containing APOE, CX3CR1, and P2RY12 were then individually knocked out (virtually) and visualized using the SCENIC+ algorithm.

**Supplementary Figure 2 Expression profile shifts following virtual KO of APOE, CX3CR1, and P2RY12 regulating TFs in mic-activated.**
Plots boxed in red indicate those that shift expression away from the resting state. Principle component analysis was performed on the TF-by-target gene matrix. The top five principal components were used to calculate the UMAP coordinates. Gene clusters were calculated from a shared-nearest-neighbor graph using the Louvain algorithm. Linear models were used to identify TFs that significantly regulated the gene clusters. TFs for the clusters containing APOE, CX3CR1, and P2RY12 were then individually knocked out (virtually) and visualized using the SCENIC+ algorithm.

**Supplementary Figure 3 Full astro-resting co-regulatory network UMAP representation.**
This pairs with Figure 3D and Supplementary Figure 4.

**Supplementary Figure 4 Detailed UMAP representation of gene regulatory networks and APOE gene cluster GO enrichment**
A-E) A UMAP visualization of each cell states' regulome that includes all pySCENIC TFs (connected to Figure 3). APOE is underlined. The TFs in red directly regulate APOE. TFs in bold black significantly upregulate the APOE gene cluster. TFs in blue are the TFs that were replicated in the ROSMAP dataset. D) Astro-resting is zoomed. The full figure can be found in Supplementary Figure 3. F) Heatmap of the GO biological process terms associated with the genes in the APOE gene clusters for each cell state. Tiles with (.) indicated an adjusted p-value < 0.05.

**Supplementary Figure 5 AD GWAS genes in the co-regulatory networks.**
A global UMAP visualization of each cell states' regulome (same as Figure 3) that highlights the GWAS genes in the same cluster with APOE (black labels) and all other GWAS genes (blue labels). Point color indicates cluster membership. The background shade of green depicts the density of AD GWAS genes in that location, where the darker the green the greater colocalization of GWAS genes in the network. Principle component analysis was performed on the TF-by-target gene matrix. The top five principal components were used to calculate the UMAP coordinates.

**Supplementary Figure 6 Other AD GWAS genes are coregulated with APOE Genes in AD GWAS loci full figures.**
(A,B) co-regulated by APOE-associated TFs and (C,D) co-clustered with APOE in co-regulation networks. Rows are colored by loci. Some genes are within two loci and therefore have two colors. These plots are the non-truncated versions of plots in Figure 5.

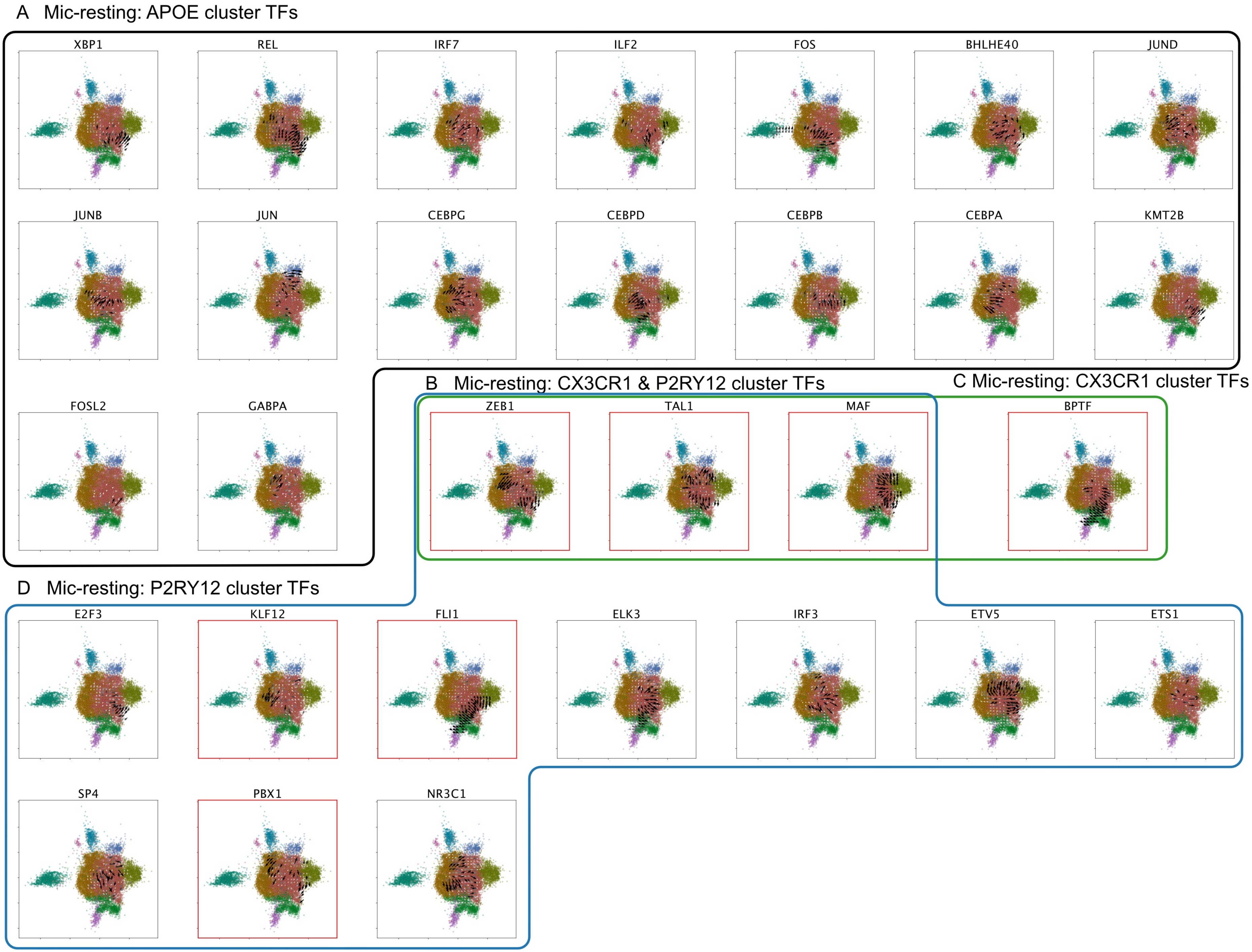
A Mic-resting: APOE cluster TFs
XBP1
REL
IRF7
ILF2
FOS
BHLHE40
JUND
JUNB
JUN
CEBPG
CEBPD
CEBPB
CEBPA
KMT2B
FOSL2
GABPA
B Mic-resting: CX3CR1 & P2RY12 cluster TFs
ZEB1
TAL1
MAF
C Mic-resting: CX3CR1 cluster TFs
BPTF
D Mic-resting: P2RY12 cluster TFs
E2F3
KLF12
FLI1
ELK3
IRF3
ETV5
ETS1
SP4
PBX1
NR3C1

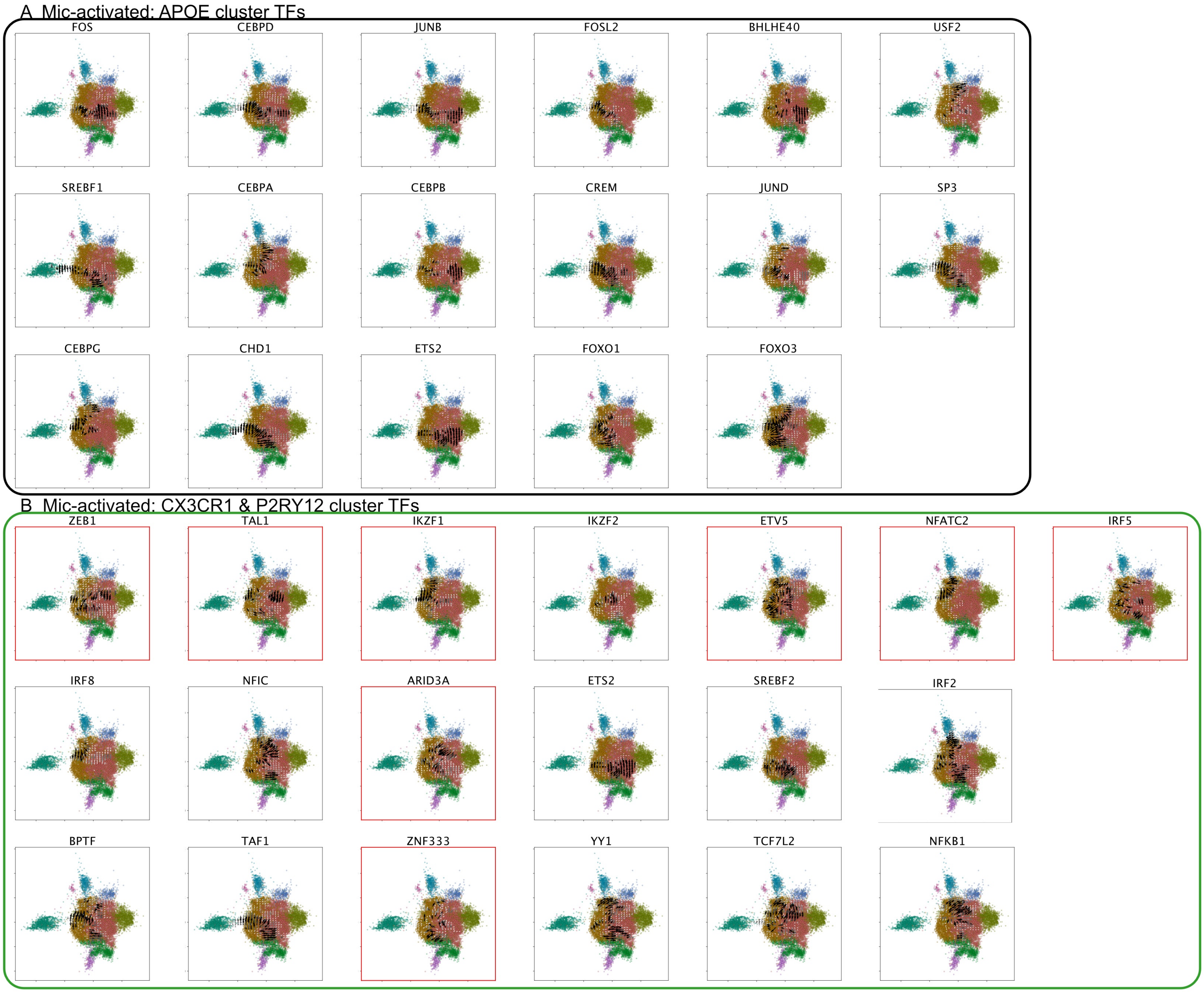
A Mic-activated: APOE cluster TFs
FOS
CEBPD
JUNB
FOSL2
BHLHE40
USF2
SREBF1
CEBPA
CEBPB
CREM
JUND
SP3
CEBPG
CHD1
ETS2
FOXO1
FOXO3
B Mic-activated: CX3CR1 & P2RY12 cluster TFs
ZEB1
TAL1
IKZF1
IKZF2
ETV5
NFATC2
IRF5
IRF8
NFIC
ARID3A
ETS2
SREBF2
IRF2
BPTF
TAF1
ZNF333
YY1
TCF7L2
NFKB1

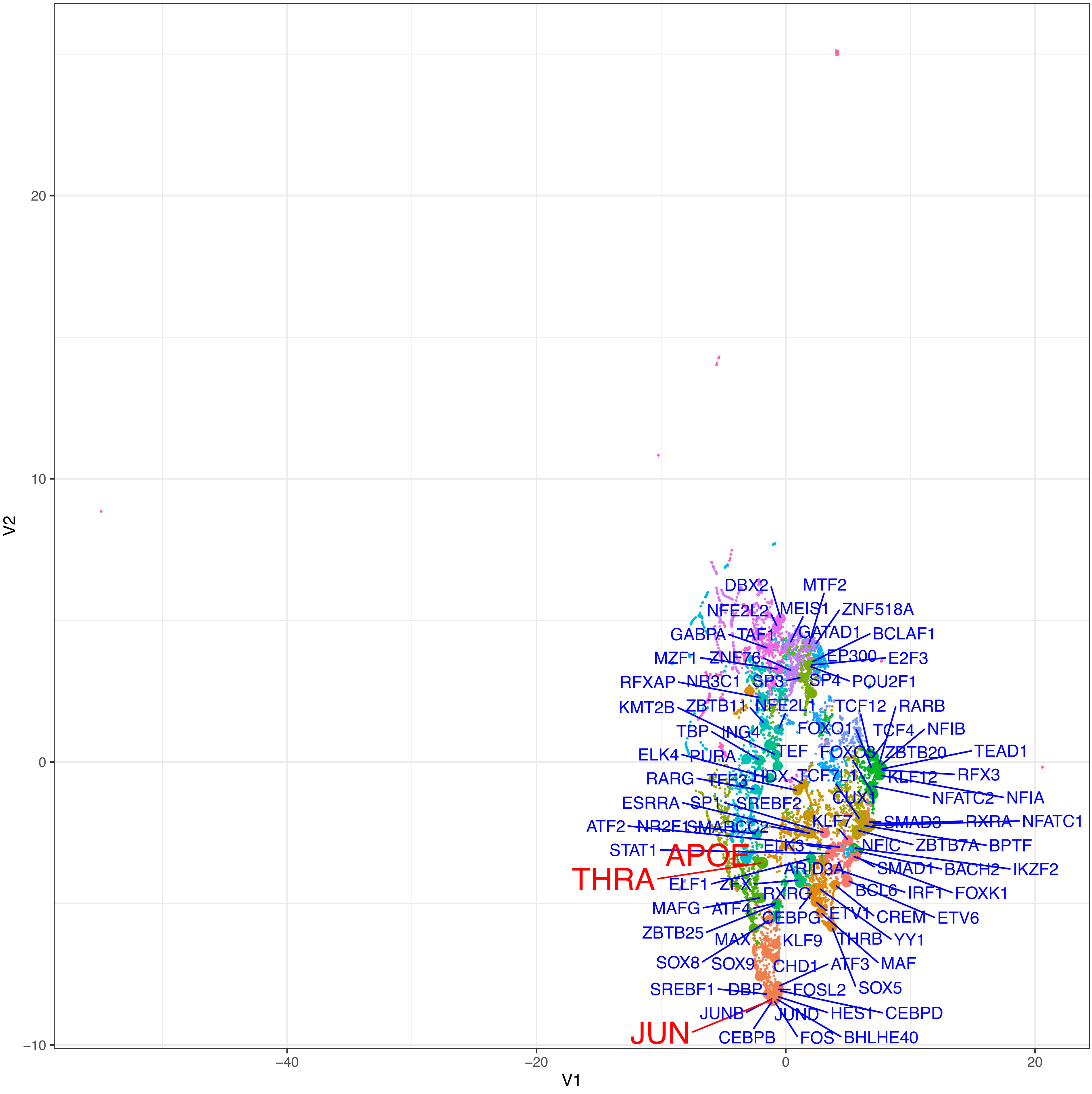
V1
V2
20
10
0
-10
-40
-20
0
20
DBX2
MTF2
NFE2L2
MEIS1
ZNF518A
GABPA
TAF1
GATAD1
BCLAF1
MZF1
ZNF76
EP300
E2F3
RFXAP
NR3C1
SP3
SP4
POU2F1
KMT2B
ZBTB11
NFE2L1
TCF12
RARB
TBP
ING4
FOXO1
TCF4
NFIB
ELK4
PURA
TEF
FOXO3
ZBTB20
TEAD1
RARG
TFE3
HDX
TCF7L1
KLF12
RFX3
ESRRA
SP1
SREBF2
CUX1
NFATC2
NFIA
ATF2
NR2F1
SMARCC2
KLF7
SMAD3
RXRA
NFATC1
STAT1
APOE
ELK3
NFIC
ZBTB7A
BPTF
THRA
ARID3A
SMAD1
BACH2
IKZF2
ELF1
ZFX
BCL6
IRF1
FOXK1
MAFG
ATF4
RXRG
CREM
ETV6
ZBTB25
CEBPG
ETV1
MAX
KLF9
THRB
YY1
SOX8
SOX9
CHD1
ATF3
MAF
SREBF1
DBP
FOSL2
SOX5
JUNB
JUND
HES1
CEBPD
JUN
CEBPB
FOS
BHLHE40

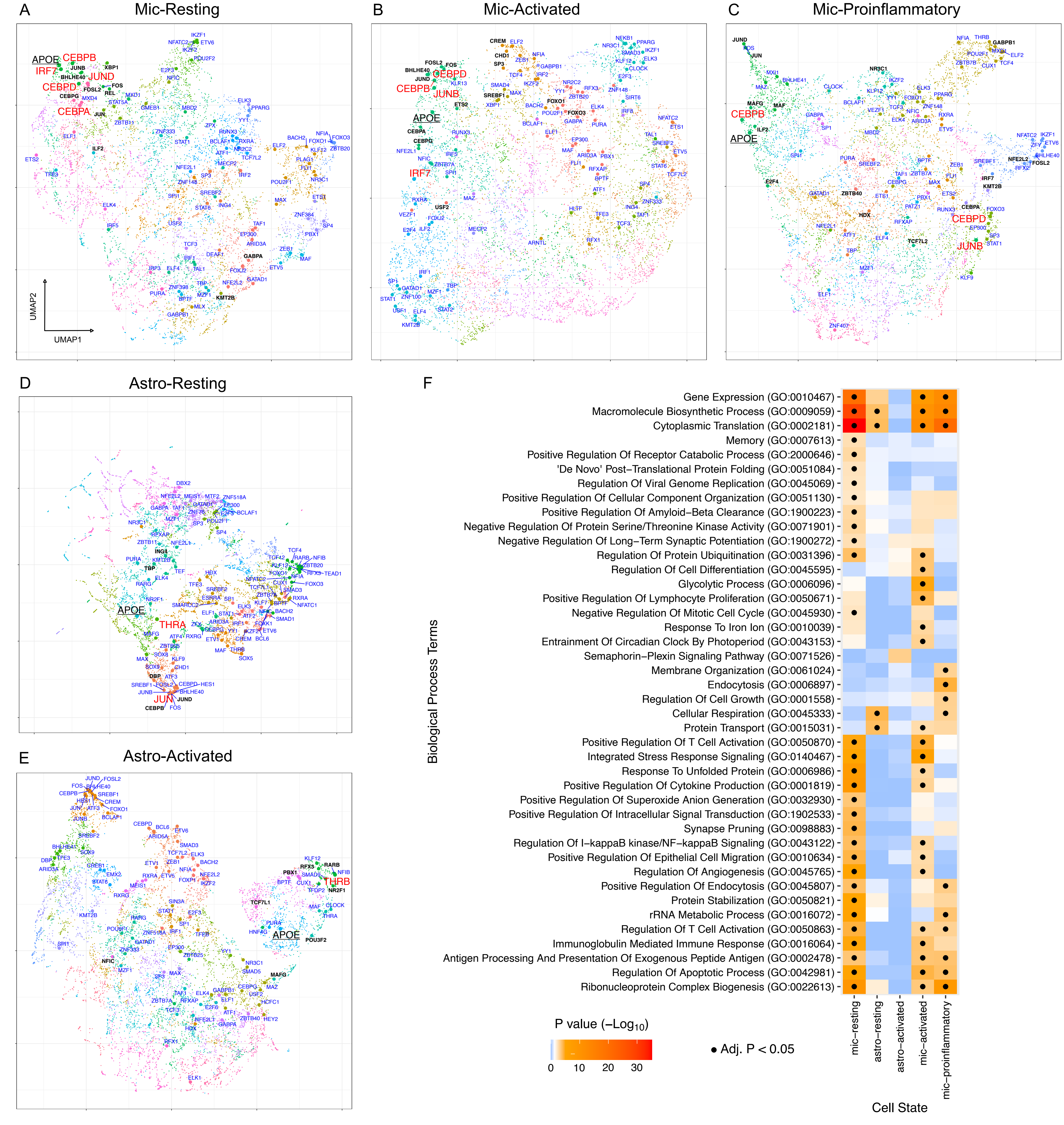
A
Mic-Resting
APOE
CEBPB
IRF7
JUND
CEBPD
CEBPA
UMAP2
UMAP1
B
Mic-Activated
CEBPD
CEBPB
JUNB
APOE
IRF7
C
Mic-Proinflammatory
CEBPB
APOE
CEBPD
JUNB
D
Astro-Resting
APOE
THRA
JUN
E
Astro-Activated
THRB
APOE
F
Gene Expression (GO:0010467)
Macromolecule Biosynthetic Process (GO:0009059)
Cytoplasmic Translation (GO:0002181)
Memory (GO:0007613)
Positive Regulation Of Receptor Catabolic Process (GO:2000646)
'De Novo' Post−Translational Protein Folding (GO:0051084)
Regulation Of Viral Genome Replication (GO:0045069)
Positive Regulation Of Cellular Component Organization (GO:0051130)
Positive Regulation Of Amyloid−Beta Clearance (GO:1900223)
Negative Regulation Of Protein Serine/Threonine Kinase Activity (GO:0071901)
Negative Regulation Of Long−Term Synaptic Potentiation (GO:1900272)
Regulation Of Protein Ubiquitination (GO:0031396)
Regulation Of Cell Differentiation (GO:0045595)
Glycolytic Process (GO:0006096)
Positive Regulation Of Lymphocyte Proliferation (GO:0050671)
Negative Regulation Of Mitotic Cell Cycle (GO:0045930)
Response To Iron Ion (GO:0010039)
Entrainment Of Circadian Clock By Photoperiod (GO:0043153)
Semaphorin−Plexin Signaling Pathway (GO:0071526)
Membrane Organization (GO:0061024)
Endocytosis (GO:0006897)
Regulation Of Cell Growth (GO:0001558)
Cellular Respiration (GO:0045333)
Protein Transport (GO:0015031)
Positive Regulation Of T Cell Activation (GO:0050870)
Integrated Stress Response Signaling (GO:0140467)
Response To Unfolded Protein (GO:0006986)
Positive Regulation Of Cytokine Production (GO:0001819)
Positive Regulation Of Superoxide Anion Generation (GO:0032930)
Positive Regulation Of Intracellular Signal Transduction (GO:1902533)
Synapse Pruning (GO:0098883)
Regulation Of I−kappaB kinase/NF−kappaB Signaling (GO:0043122)
Positive Regulation Of Epithelial Cell Migration (GO:0010634)
Regulation Of Angiogenesis (GO:0045765)
Positive Regulation Of Endocytosis (GO:0045807)
Protein Stabilization (GO:0050821)
rRNA Metabolic Process (GO:0016072)
Regulation Of T Cell Activation (GO:0050863)
Immunoglobulin Mediated Immune Response (GO:0016064)
Antigen Processing And Presentation Of Exogenous Peptide Antigen (GO:0002478)
Regulation Of Apoptotic Process (GO:0042981)
Ribonucleoprotein Complex Biogenesis (GO:0022613)
Biological Process Terms
mic−resting
astro−resting
astro−activated
mic−activated
mic−proinflammatory
Cell State
P value (−Log10)
0
10
20
30
• Adj. P < 0.05

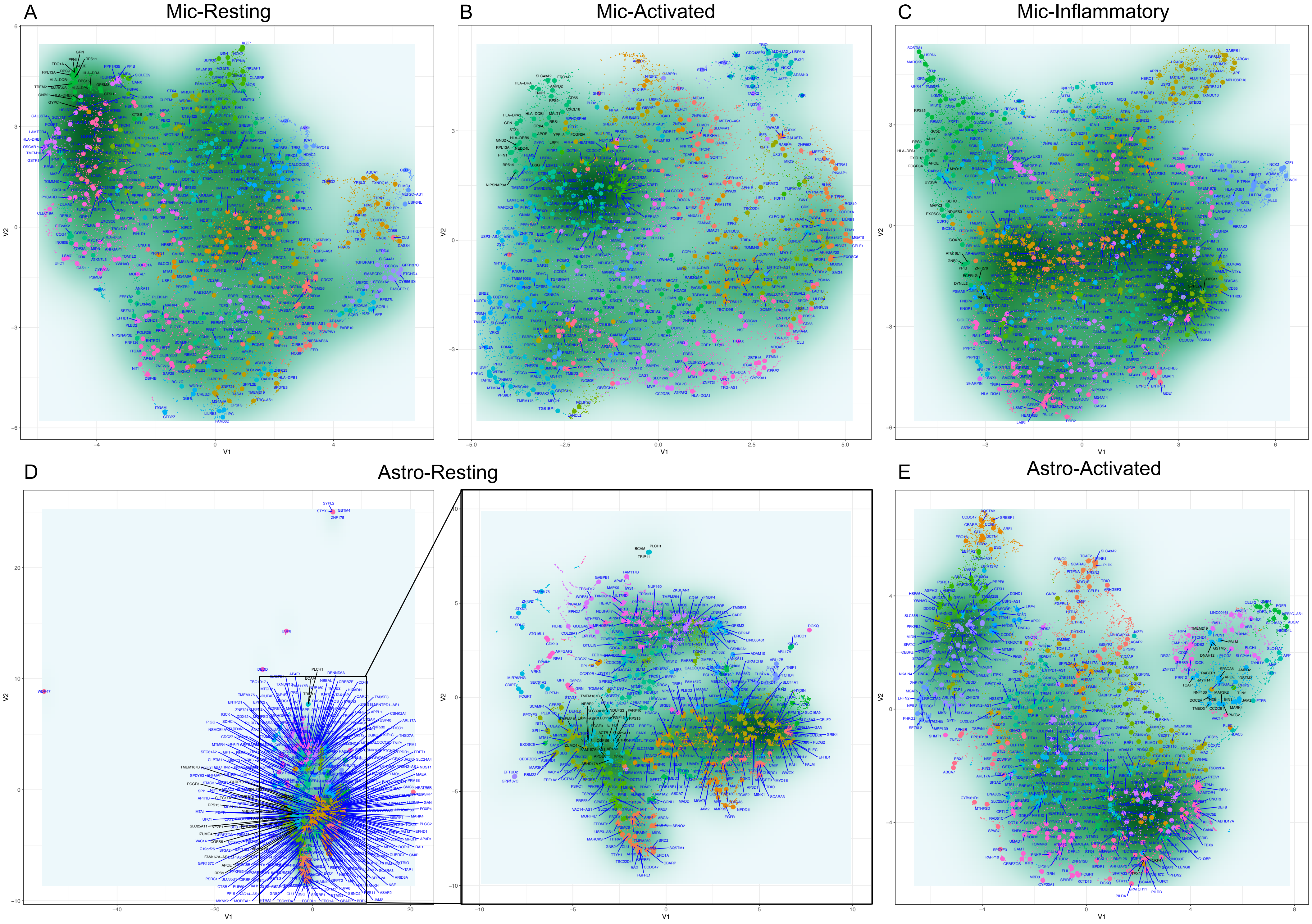
A
Mic-Resting
B
Mic-Activated
C
Mic-Inflammatory
D
Astro-Resting
E
Astro-Activated
V1
V2

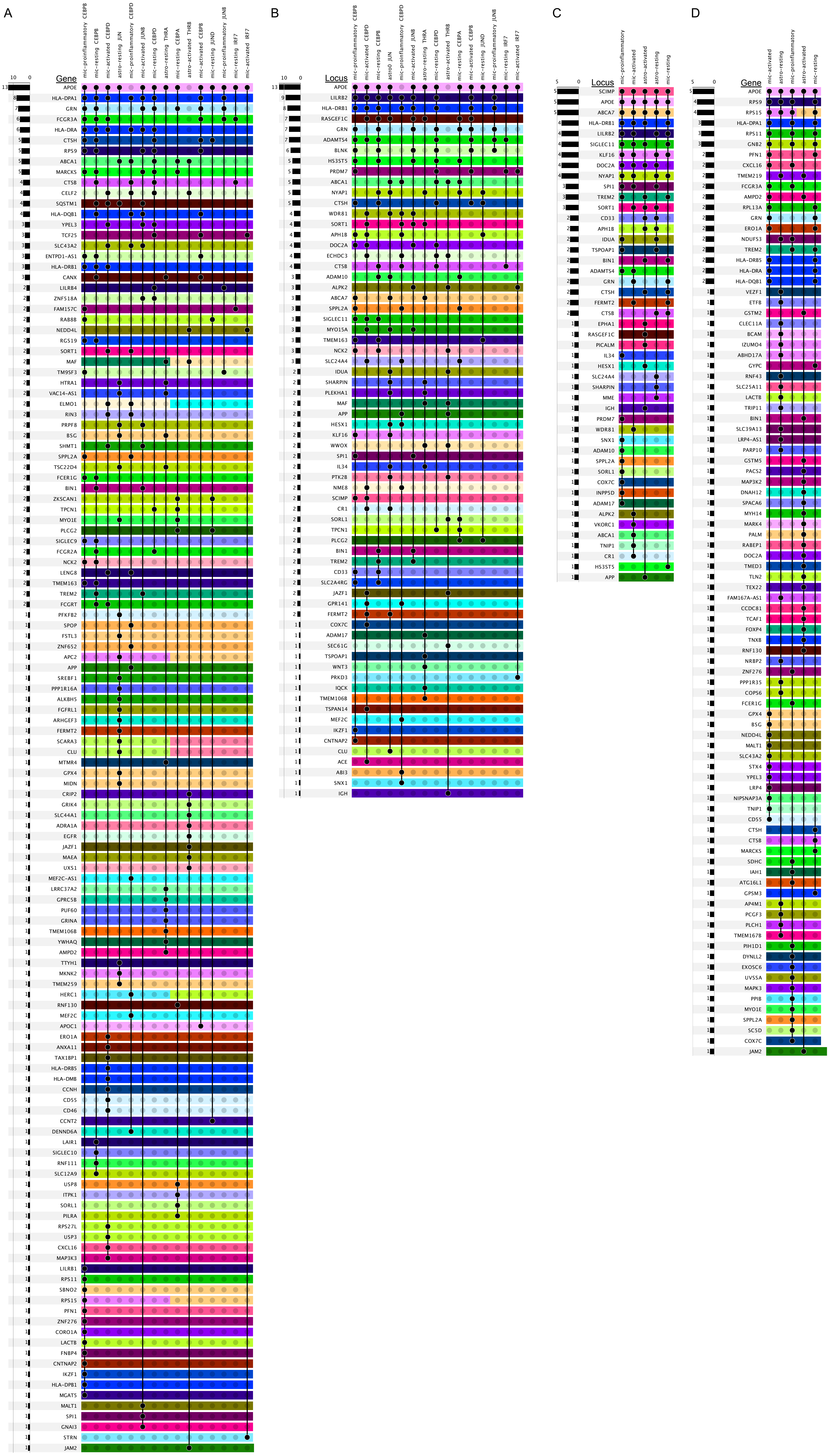

A
mic-proinflammatory CEBPB
mic-resting CEBPB
mic-activated CEBPD
astro-resting JUN
mic-proinflammatory CEBPD
mic-activated JUNB
mic-resting CEBPD
astro-resting THRA
mic-resting CEBPA
astro-activated THRB
mic-activated CEBPB
mic-resting JUND
mic-proinflammatory JUNB
mic-resting IRF7
mic-activated IRF7
Gene
APOE
HLA-DPA1
GRN
FCGR3A
HLA-DRA
CTSH
RPS9
ABCA1
MARCKS
CTSB
CELF2
SQSTM1
HLA-DQB1
YPEL3
TCF25
SLC43A2
ENTPD1-AS1
HLA-DRB1
CANX
LILRB4
ZNF518A
FAM157C
RAB8B
NEDD4L
RGS19
SORT1
MAF
TM9SF3
HTRA1
VAC14-AS1
ELMO1
RIN3
PRPF8
BSG
SHMT1
SPPL2A
TSC22D4
FCER1G
BIN1
ZKSCAN1
TPCN1
MYO1E
PLCG2
SIGLEC9
FCGR2A
NCK2
LENG8
TMEM163
TREM2
FCGRT
PFKFB2
SPOP
FSTL3
ZNF652
APC2
APP
SREBF1
PPP1R16A
ALKBH5
FGFRL1
ARHGEF3
FERMT2
SCARA3
CLU
MTMR4
GPX4
MIDN
CRIP2
GRIK4
SLC44A1
ADRA1A
EGFR
JAZF1
MAEA
UXS1
MEF2C-AS1
LRRC37A2
GPRC5B
PUF60
GRINA
TMEM106B
YWHAQ
AMPD2
TTYH1
MKNK2
TMEM259
HERC1
RNF130
MEF2C
APOC1
ERO1A
ANXA11
TAX1BP1
HLA-DRB5
HLA-DMB
CCNH
CD55
CD46
CCNT2
DENND6A
LAIR1
SIGLEC10
RNF111
SLC12A9
USP8
ITPK1
SORL1
PILRA
RPS27L
USP3
CXCL16
MAP3K3
LILRB1
RPS11
SBNO2
RPS15
PFN1
ZNF276
CORO1A
LACTB
FNBP4
CNTNAP2
IKZF1
HLA-DPB1
MGAT5
MALT1
SPI1
GNAI3
STRN
JAM2
B
mic-proinflammatory CEBPB
mic-activated CEBPD
mic-resting CEBPB
astro0 JUN
mic-proinflammatory CEBPD
mic-activated JUNB
astro-resting THRA
mic-resting CEBPD
astro-activated THRB
mic-resting CEBPA
mic-activated CEBPB
mic-resting JUND
mic-proinflammatory JUNB
mic-resting IRF7
mic-activated IRF7
Locus
APOE
LILRB2
HLA-DRB1
RASGEF1C
GRN
ADAMTS4
BLNK
HS3ST5
PRDM7
ABCA1
NYAP1
CTSH
WDR81
SORT1
APH1B
DOC2A
ECHDC3
CTSB
ADAM10
ALPK2
ABCA7
SPPL2A
SIGLEC11
MYO15A
TMEM163
NCK2
SLC24A4
IDUA
SHARPIN
PLEKHA1
MAF
APP
HESX1
KLF16
WWOX
SPI1
IL34
PTK2B
NME8
SCIMP
CR1
SORL1
TPCN1
PLCG2
BIN1
TREM2
CD33
SLC2A4RG
JAZF1
GPR141
FERMT2
COX7C
ADAM17
SEC61G
TSPOAP1
WNT3
PRKD3
IQCK
TMEM106B
TSPAN14
MEF2C
IKZF1
CNTNAP2
CLU
ACE
ABI3
SNX1
IGH
C
mic-proinflammatory
mic-activated
astro-activated
astro-resting
mic-resting
Locus
SCIMP
APOE
ABCA7
HLA-DRB1
LILRB2
SIGLEC11
KLF16
DOC2A
NYAP1
SPI1
TREM2
SORT1
CD33
APH1B
IDUA
TSPOAP1
BIN1
ADAMTS4
GRN
CTSH
FERMT2
CTSB
EPHA1
RASGEF1C
PICALM
IL34
HESX1
SLC24A4
SHARPIN
MME
IGH
PRDM7
WDR81
SNX1
ADAM10
SPPL2A
SORL1
COX7C
INPP5D
ADAM17
ALPK2
VKORC1
ABCA1
TNIP1
CR1
HS3ST5
APP
D
mic-activated
astro-resting
mic-proinflammatory
astro-activated
mic-resting
Gene
APOE
RPS9
RPS15
HLA-DPA1
RPS11
GNB2
PFN1
CXCL16
TMEM219
FCGR3A
AMPD2
RPL13A
GRN
ERO1A
NDUFS3
TREM2
HLA-DRB5
HLA-DRA
HLA-DQB1
VEZF1
ETFB
GSTM2
CLEC11A
BCAM
IZUMO4
ABHD17A
GYPC
RNF43
SLC25A11
LACTB
TRIP11
BIN1
SLC39A13
LRP4-AS1
PARP10
GSTM5
PACS2
MAP3K2
DNAH12
SPACA6
MYH14
MARK4
PALM
RABEP1
DOC2A
TMED3
TLN2
TEX22
FAM167A-AS1
CCDC81
TCAF1
FOXP4
TNXB
RNF130
NRBP2
ZNF276
PPP1R35
COPS6
FCER1G
GPX4
BSG
NEDD4L
MALT1
SLC43A2
STX4
YPEL3
LRP4
NIPSNAP3A
TNIP1
CD55
CTSH
CTSB
MARCKS
SDHC
IAH1
ATG16L1
GPSM3
AP4M1
PCGF3
PLCH1
TMEM167B
PIH1D1
DYNLL2
EXOSC6
UVSSA
MAPK3
PPIB
MYO1E
SPPL2A
SC5D
COX7C
JAM2

### Supplementary Tables

| *Supplementary Table 1 Cell state summaries* | Mic resting | Mic activated | Mic inflammatory | Astro resting | Astro activated |
|---|---|---|---|---|---|
| TF regulators | 114 | 114 | 109 | 119 | 127 |
| ROSMAP TF regulators | 109 | 112 | 106 | 107 | 113 |
| Replicated TFs | 94 | 100 | 86 | 97 | 101 |
| Co-regulatory network clusters | 27 | 23 | 24 | 31 | 30 |
| Genes in APOE cluster | 199 | 365 | 352 | 310 | 409 |
| TFs regulating APOE cluster | 16 | 17 | 18 | 6 | 9 |

*Supplementary Table 2 TFs for each cell state*
*https://wustl.box.com/v/pySCENIC-TF-intersect-upset*

*Supplementary Table 3 APOE gene cluster gene set enrichment analysis results*
*https://wustl.box.com/v/APOE-gene-cluster-enrichment*

*Supplementary Table 4 State-specific TF gene set enrichment analysis results*
*https://wustl.box.com/v/TF-enrichr-results*

*Supplementary Table 5 Genes co-regulated with APOE*
*https://wustl.box.com/v/genes-coregulated-with-APOE*

*Supplementary Table 6 Genes in AD GWAS loci*
*https://wustl.box.com/v/genes-in-AD-GWAS-loci*
